# Diverse Monogenic Subforms of Human Spermatogenic Failure

**DOI:** 10.1101/2022.07.19.22271581

**Authors:** Liina Nagirnaja, Alexandra M. Lopes, Wu-Lin Charng, Brian Miller, Rytis Stakaitis, Ieva Golubickaite, Alexandra Stendahl, Tianpengcheng Luan, Corinna Friedrich, Eisa Mahyari, Eloise Fadial, Laura Kasak, Katinka Vigh-Conrad, Manon S. Oud, Miguel J. Xavier, Samuel R. Cheers, Emma R. James, Jingtao Guo, Timothy G Jenkins, Antoni Riera-Escamilla, Alberto Barros, Filipa Carvalho, Susana Fernandes, João Gonçalves, Christina A. Gurnett, Niels Jørgensen, Davor Jezek, Emily S Jungheim, Sabine Kliesch, Robert I. McLachlan, Kenan R Omurtag, Adrian Pilatz, Jay Sandlow, James Smith, Michael L. Eisenberg, James M Hotaling, Keith A. Jarvi, Margus Punab, Ewa Rajpert-De Meyts, Douglas T. Carrell, Csilla Krausz, Maris Laan, Moira K. O’Bryan, Peter N. Schlegel, Frank Tüttelmann, Joris A. Veltman, Kristian Almstrup, Kenneth I. Aston, Donald F. Conrad

## Abstract

Non-obstructive azoospermia (NOA) is the most severe form of male infertility and typically incurable with current medicine. Due to the biological complexity of sperm production, defining the genetic basis of NOA has proven challenging, and to date, the most advanced classification of NOA subforms is based on simple description of testis histology. In this study, we exome-sequenced over 1,000 clinically diagnosed NOA cases and identified a plausible recessive Mendelian cause in 20%. Population-based testing against fertile controls identified 27 genes as significantly associated with azoospermia. The disrupted genes are primarily on the autosomes, enriched for undescribed human “knockouts”, and, for the most part, have yet to be linked to a Mendelian trait. Integration with single-cell RNA sequencing of adult testes shows that, rather than affecting a single cell type or pathway, azoospermia genes can be grouped into molecular subforms with highly synchronized expression patterns, and analogs of these subforms exist in mice. This analysis framework identifies groups of genes with known roles in spermatogenesis but also reveals unrecognized subforms, such as a set of genes expressed specifically in mitotic divisions of type B spermatogonia. Our findings highlight NOA as an understudied Mendelian disorder and provide a conceptual structure for organizing the complex genetics of male infertility, which may serve as a basis for disease classification more advanced than histology.

## Introduction

Non-obstructive azoospermia, or lack of sperm in the ejaculate due to disruption of spermatogenesis, is a multifactorial trait with a prevalence of 0.4%-2% in the male population^1–3^ and has an incidence of around 10% in cohorts of infertile men^4^. Mounting evidence suggests that male infertility is broadly associated with late-onset somatic comorbidities, including cancer, cardiovascular disease, and other chronic diseases^5^. The mechanisms by which these risks are linked are generally unknown. From the perspective of patient counseling, disease prevention and management, it is thus becoming increasingly relevant to establish the genetic origins of male infertility and spermatogenic impairment. During pioneering genetic investigations over 40 years ago, large Y chromosome deletions were identified as a highly penetrant genetic cause of human azoospermia^6^. The advent of next generation sequencing has renewed the hunt for Mendelian forms of azoospermia and the number of confirmed monogenic causes has since grown to encompass at least 40 loci^7, 8^. Nevertheless, our understanding of genetic causes of male infertility phenotypes is still lacking considering that the number of genes linked to the trait in mice is over 500^9^, half of which are specifically implicated in azoospermia^10^. The few genomewide studies published have shown promise of using whole-exome and whole-genome sequencing to identify the missing genetic causes of azoospermia^11–15^. What is needed now is a conceptual framework to organize what appears to be a very large collection of monogenic disorders that we collectively call “male infertility”. We hypothesized that distinct subforms of NOA exist, currently invisible by standard histological classification, but detectable at a molecular level. In this study, we test this hypothesis through analysis of the largest collection of azoospermia cases to date, combined with a new single-cell RNA sequencing (scRNA-seq) atlas of human testis gene expression (Fig. 1). The results presented provide a broad and detailed survey of rare genetic changes causing azoospermia in humans, and a framework for organizing these changes into subgroups, for improved characterization and personalized management of infertility patients.

**Fig. 1.**
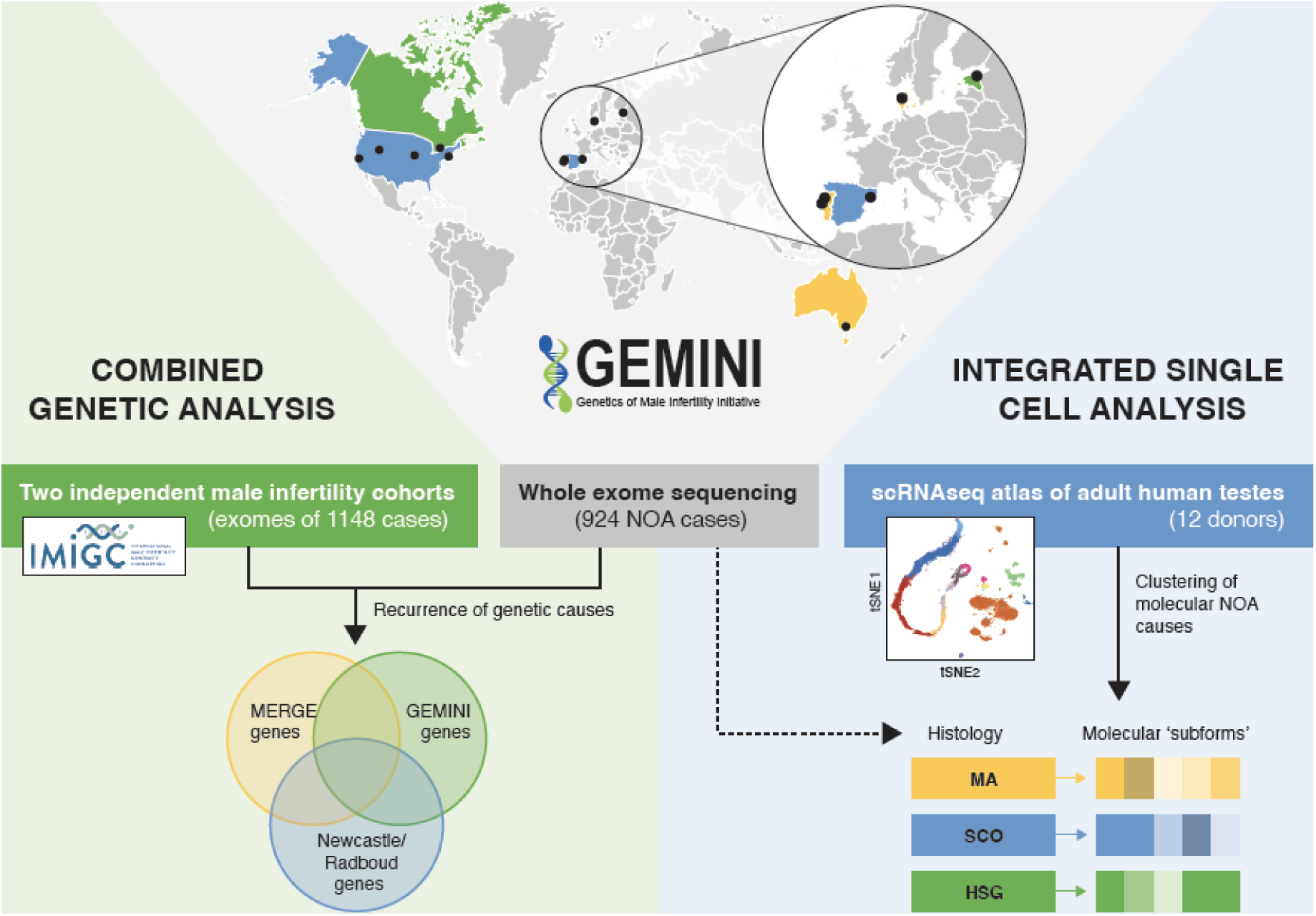
Overview of the study. DNA samples from NOA cases recruited in 11 centers across the world underwent WES to detect rare recessive variants underlying the disease. The identified Mendelian causes of NOA were additionally screened for in independent male infertility cohorts to map recurrent causes of NOA. An integrated analysis with scRNAseq atlas of adult human testis enabled to cluster the molecular NOA causes into ‘subforms’ that go beyond the visual classification of the disease based on histological findings.

## Results

### Variation prioritization identifies potential monogenic causes in 20% of NOA cases

Through the GEnetics of Male INfertility Initiative (GEMINI), we performed whole exome sequencing (WES) on 1,011 unrelated men diagnosed with spermatogenic failure, the vast majority with unexplained NOA. Carriers of known genetic causes of azoospermia or large structural variants were excluded using the processed sequencing dataset (Extended Data Fig. 1, Supplementary Discussion). The final GEMINI cohort analyzed here consisted of 924 unrelated cases recruited in eleven centers across seven countries (Fig. 1, Extended Data Figs. 1b, 2, Supplementary Discussion, Supplementary Table 1). Testis biopsies were performed on 42% of the subjects. These biopsies were used to classify patients into three distinct histological subtypes-Sertoli Cell Only (SCO), complete lack of germ cells and the most prevalent diagnosis (n=248 men), followed by maturation arrest (MA, arrest of spermatogenesis, n=101) and hypospermatogenesis (HSG, n=37), where the spermatogenic output is severely reduced due to significant variation in inter-tubule content, with or without complete spermatogenesis (Extended Data Fig. 2).

We prioritized rare (minor allele frequency <1% in gnomAD) likely damaging lesions compatible with recessive Mendelian disease models by using an analysis pipeline based on Population Sampling Probability (PSAP, Methods, Fig 2a, Extended Data Fig. 3, Supplementary Table 2) ^16^. The prioritized variants were further filtered by taking into account the mutation type and the testicular expression patterns of the underlying genes (Methods) yielding a total of 312 variants most likely to have an impact on spermatogenesis. Experimental validation confirmed 98% (n=82/84) of SNVs and 93% (n=38/41) of INDELs tested, whereas a decreased rate was detected for CNVs (43%, n=3/7) as expected due to the lower accuracy of detecting structural rearrangements from exome data ^17^.

**Fig. 2.**
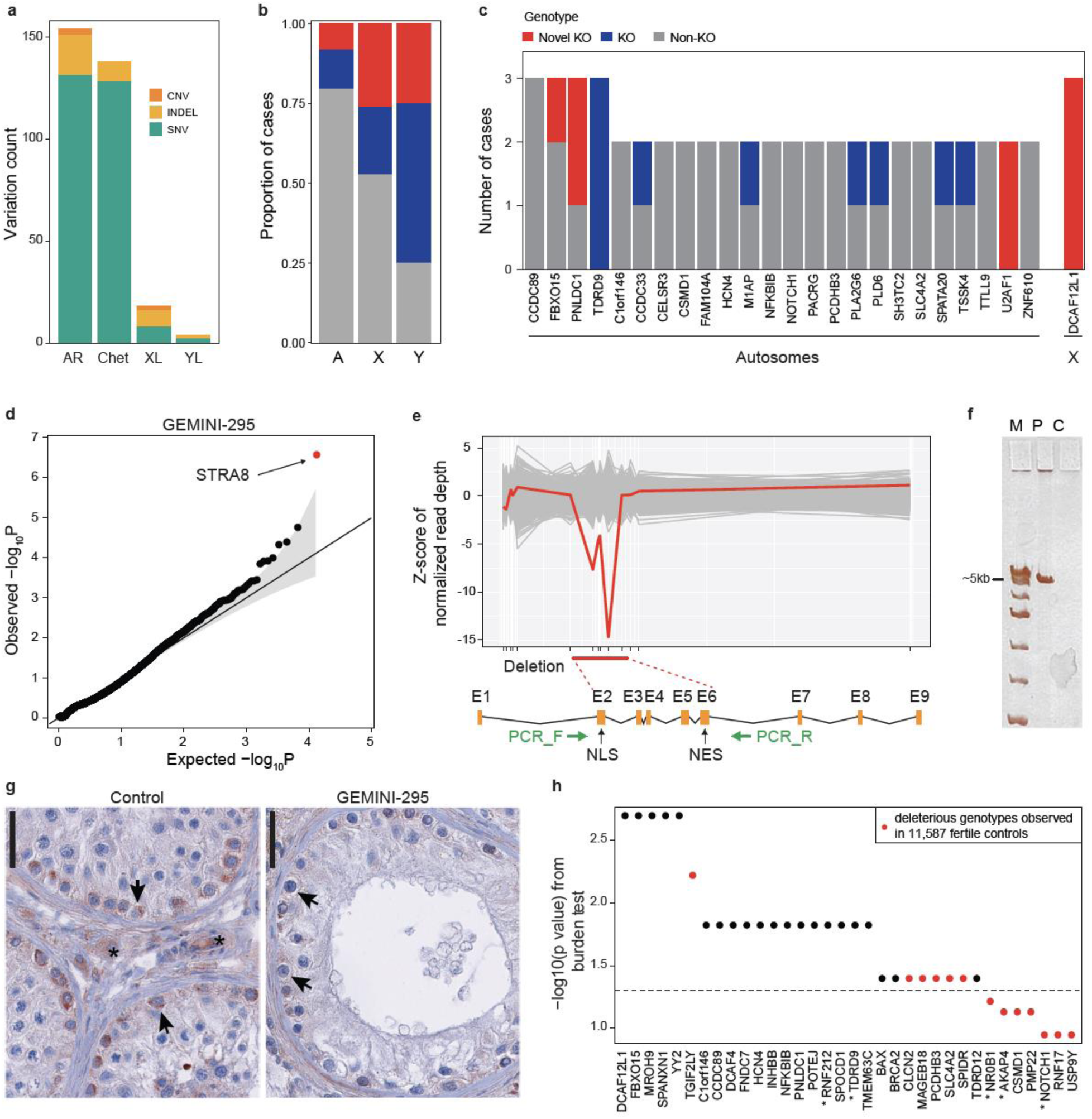
Variant prioritization and burden testing in NOA cases. **a**, Distribution of the prioritized variation types across inheritance modes. AR, autosomal recessive, Chet, Compound heterozygous, XL, X-linked, YL, Y-linked. **b**, Predicted hemizygous loss-of-function genotypes appear to be enriched on sex chromosomes compared to biallelic loss-of-function genotypes on autosomes (A). *p<0.05. **c**, Summary of all genes with multiple case findings in the GEMINI cohort. **d**, PSAP p-values of all variants detected in patient GEMINI-295, the carrier of the biallelic *STRA8* deletion. *STRA8* CNV (the smallest p-value) was prioritized as the most likely cause of NOA in GEMINI-295. **e**, Z-scores of normalized read depth of exome sequencing data around the *STRA8* locus, plotted against the *STRA8* gene model. NLS; nuclear localization; NES, nuclear export signal. Green arrows, PCR primers. **f,** PCR spanning the predicted deletion region yielded a short ∼5 kb product in GEMINI-295 (P) validating the homozygous *STRA8* deletion. C, control; M, ladder. **g**, In control testis, clear staining of STRA8 was observed in spermatogonia (arrows), together with background staining in peritubular and interstitial cells (asterisk). In GEMINI-295, STRA8 protein staining in spermatogonia is much lower (arrows), presumably due to nonsense mediated decay of the transcript. Bar represents 10*μ*m. **h,** Burden test results from comparison of a combined cohort of 2,072 NOA cases with 11,587 fertile controls. Thirty-four genes with prioritized variation in the GEMINI cohort were selected for burden testing; of these 27 were nominally associated with NOA (Methods). * indicates genes with at least “limited” clinical validity defined by ^7^.

Using PSAP prioritization, we identified a potential molecular cause of the disease for 19.3% (n=178/924) of the NOA cases, when considering the SNVs and INDELs only. The PSAP pipeline is a filtering tool; some variants that are completely unrelated to infertility may pass this filter. In order to estimate what fraction of prioritized variants are false positives, we also applied PSAP to an institutional control cohort of 2,665 individuals with non-reproductive disorders (Extended Data Table 1). Only 6% of controls carried prioritized variants, indicating that the discovery rate in GEMINI is much higher than background (P=1.7x10^-27^), but also indicating that as many as ⅓ of prioritized variants in GEMINI may be false positives.

The NOA cohort included 72 unrelated consanguineous cases that were expected to carry damaging recessive variation leading to the disease ^8^. Consistently, a possible genetic cause of NOA was identified in 76.4% of the consanguineous men, significantly higher than the remainder of the cohort 14.9%; Fisher’s exact test P=8.3x10^-28^; Supplementary Discussion).

Additionally, two unrelated NOA cases from Utah were identified with uniparental isodisomy of chromosome 2 (UPD2) and chromosome 4 (UPD4), and with prioritized variation in *INHBB* ^18^ (regulation of FSH production) and *POLN* (involved in recombination) on affected chromosomes, respectively (Supplementary Discussion; Extended Data Fig. 4a). The overall detection rate of a candidate NOA lesion with all variation types combined reached 20%, an estimate similar to previously reported rates in genomic studies of male infertility ^14, 15^.

### Population-based statistical tests associate 27 genes with azoospermia

The PSAP analysis described above is useful as a prioritization filter that is sensitive to monogenic disease mutations in n=1 cases, but it is not a replacement for association testing, which remains a gold standard for definitive identification of disease genes. To perform association testing, we assembled two independent cohorts of infertile men from MERGE (n=817 cases from Germany) and Newcastle/Radboud studies (total n=331; n=286 from Netherlands, n=45 UK), as well as a large control cohort of 11,587 fertile parents (Fig. 1, Methods). Of the 221 putative disease genes prioritized in the GEMINI cohort, 18.9% were disrupted in at least one other NOA cohort (Supplementary Table 3). Three genes were affected in all NOA cohorts: *M1AP,* recently published in a cross-center report as a novel cause of autosomal recessive meiotic arrest in men, and the uncharacterized genes *KCTD19* and *YY2* (Supplementary Table 3)^19, 20^.

Using the combined NOA cohorts (n=2,072 cases) and 11,587 fertile controls, we performed burden testing on 34 genes with prioritized mutations in at least 2 cases (Fig. 2h). We reasoned that only testing genes with multiple prioritized variants would limit multiple testing and maximize power. To increase rigor, we restricted our analysis to only genotypes compatible with recessive disease (e.g. homozygous and hemizygous changes). Due to data use limitations we were unable to obtain data on compound heterozygous changes from controls, thus limiting some of the genes that could be tested (Methods). Burden testing identified 27 genes with significant p-values, providing strong evidence that variation in these genes is likely deleterious for human male infertility (Supplementary Table 4). Especially strong evidence was found for 21 genes, for which we identified multiple highly deleterious genotypes in cases, and none in 11,587 fertile controls (Fig 2h). Of these, only *TDRD9* and *RNF212* were identified as having at least “limited” clinical validity in a recent review of azoospermia research ^7^.

### Undescribed human “knock-outs” in azoospermia

We next identified rare human biallelic loss-of-function variants, or “knock-outs” (KO), enrichment of which has been observed for Mendelian disease, developmental disorders, and autism ^21^. A high-confidence complete KO was predicted for 50/221 NOA genes identified in this study with an enrichment on X and Y chromosomes (p=0.018 and p=0.032 vs autosomes, respectively; hypergeometric test, Fig. 2b) that have historically been considered the main source of azoospermia defects. On chromosome X, 9/19 (47%) genotypes were KO, including in genes *MAGEA3*, *MAGEB18* and *MAGEC3*, the members of the cancer/testis-antigen family modulating reproductive success ^22^. On chromosome Y, prioritized LoF mutations were found in *USP9Y, DDX3Y, TGIF2LY*, and *ZFY*, the latter three yet to be linked to male infertility in humans.

Notably, 38.0% (n=19) of the KO genes represented the first instances of human KO observed across more than 224,000 individuals in five large human variation datasets, including gnomAD and the Human Gene Mutation Database (Methods; Supplementary Table 6). For comparison, significantly fewer cases of novel predicted KOs were observed among the 2265 institutional controls considering only autosomal genes (n=11; one-tailed binomial test, P=5.4x10^-7^). Three genes were recurrently affected by biallelic or hemizygous loss-of-function variants, including poorly studied X-linked *DCAF12L1* (Fig. 2c).

In addition to LoF variants, we prioritized five CNVs (1.8 kb duplication and five homozygous/hemizygous deletion events of 0.48 kb-1.1 Mb) that are expected to abolish the function of the underlying genes (Extended Data Fig. 5a; Supplementary Discussion). The rearranged genes were novel in the context of male infertility, with an exception of *STRA8* where a homozygous splice variant was recently found in an NOA/severe oligozoospermia cohort ^15^. We identified and validated a homozygous ∼6kb deletion in the gene, which was predicted as the top candidate cause of NOA in the patient (Fig. 2d-f, Extended Data Figs. 5b-d). STRA8 is considered a meiotic gatekeeper and plays a vital role in meiotic initiation upon induction by retinoic acid in both female and male germline ^23^. The NOA patient with the *STRA8* deletion displayed a maturation arrest phenotype where pre-pachytene spermatocytes were rarely observed and germ cells beyond the pre-pachytene stage were not detected, largely consistent with the *Stra8*^-/-^ mouse model (Fig. 2g; Extended Data Fig. 6) ^23^. These data indicate that undescribed human KOs contribute to the loss of fertility in men.

### Clinical validity assessment and the role of recurrent gene mutations in establishing pathogenicity

Although the aim of this study was to explore and compile an extensive catalog of recessive variation possibly leading to NOA among unrelated cases, we evaluated the clinical validity of the variants to identify the genes with the best prospects for clinical testing. Potentially diagnostic variants classified as “Pathogenic” or “Likely Pathogenic” were identified in 29 genes disrupted in 37 cases from the GEMINI cohort (Supplementary Table 5) based on the guidelines from the American College of Medical Genetics and Genomics (ACMG)^24^. These genes consisted of eight with existing human genetic evidence, whereas three appear to be implicated in human or mouse male infertility for the first time based on the evidence compiled here: *DCAF12L1*, *DCAF4* and *YY2* (Supplementary Table 5).

Recurrent observations are a key for classifying novel disease variation as ‘(Likely) Pathogenic’ according to the ACMG guidelines. We observed only 25/221 (11.3%) of prioritized genes in multiple subjects, with a maximum of three recurrent hits seen for four genes (Fig. 2c; Supplementary Table 7). This is in accordance with the biological complexity of the testis where 77% of all human protein coding genes are expressed^25^. False positives identified by PSAP prioritization should only lightly influence this observation. Depending on assumptions about how false positives are distributed among genes and cases, the expected rate of recurrence ranges from 8.5%-16.1%.

For the majority of the NOA cases with a detected possible cause (77%), a single gene was identified, consistent with a monogenic form of the disease, whereas the remaining subjects carried recessive variation in up to five loci depending on the degree of consanguinity (Extended Data Fig. 4b). Based on these observations, and a simple model of NOA genetic architecture, we estimate the total number of monogenic azoospermia genes is around 625 (Fig 2a; Methods). Incorporating the estimated FDR of PSAP prioritization in the calculations, the total number drops 12% to 550. The cohort sizes required for detecting multiple occurrences of at least half of all azoospermia genes are thus an order of magnitude higher among outbred cases (n∼6700) than has been feasible until now (Fig. 3b), while the number of consanguineous cases required for equivalent power is much lower (n∼1300) (Fig. 3b). These results highlight the necessity for large cohort studies for identifying NOA disease variation with a diagnostic value as per the ACMG guidelines.

**Fig. 3.**
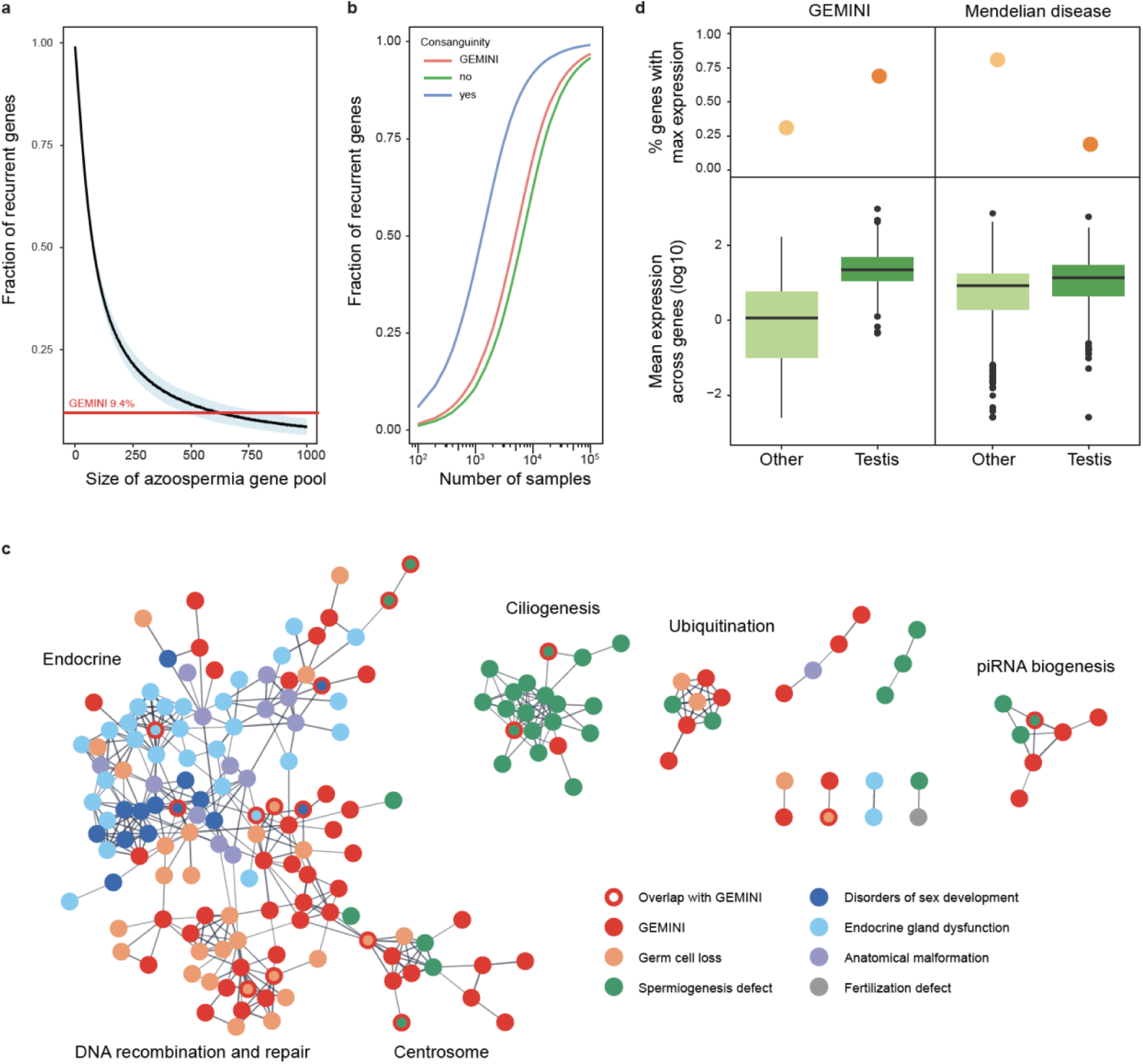
Characteristics of NOA genes in the context of male infertility and the broader Mendelian disease landscape. **a**, Estimation of the total number of azoospermia genes. To detect 9.4% of genes recurrently in non-consanguineous cases with predominantly monogenic NOA, as seen in GEMINI, a gene pool of roughly 625 azoospermia genes would be expected. Blue area represents standard deviation around the mean of 1000 iterations. **b**, Projected number of samples required for observing recurrent gene hits in NOA cohorts considering the estimated pool of 625 azoospermia genes and discovery rates of 76% for consanguineous cases, 20% for GEMINI overall (8% of consanguineous cases) and 15% for non-consanguineous cases. **c**, High confidence protein-protein interactions between GEMINI genes and known male infertility disease genes from Oud et al. 2019^7^ (Supplementary Table 9). **d**, Expression patterns of NOA genes are distinct from that of known recessive Mendelian disease genes (n=615) in the GTEx atlas of 52 human tissues. NOA genes are more likely to have maximum expression in testis (top panel) and greater testis-specific expression (lower panel). Center line, median; box limits, upper and lower quartiles; whiskers, 1.5x interquartile range; points, outliers.

### NOA as an understudied Mendelian disorder

In spite of the growing number of studies on the genetics of NOA, the total number of the genes linked to the disease has remained minuscule. Consistently, we observed a low fraction (7.7%) of genes previously reported in association with male infertility in humans (Extended Data Table 2). The remainder represent novel NOA candidate genes in men, of which at least 29.9% (61/204) were known to cause sub/infertility upon disruption in mice. A protein-protein interaction (PPI) network analysis of the prioritized genes identified numerous well defined subgroups of interacting proteins with clear roles in testis function (Fig. 3c).

To expand the context to all known Mendelian trait loci, we intersected the prioritized gene list with known Mendelian disease genes in OMIM, an online database of Mendelian traits. For the fifty-one (24.9%) NOA genes in OMIM, infertility was the most frequent reported disease type (n=13), followed by developmental disorders (n=11) and cancer (n=7) (Supplementary Table 8). Collectively these findings highlight primary testicular failure as a poorly studied Mendelian disorder. Cases of idiopathic male infertility can provide knowledge about Mendelian disease that is under-sampled in other study populations, likely due to tissue-specific functions of many NOA genes. Indeed, by analysis of expression patterns across 52 human tissues in the GTEx database^26^, we find that genes with high expression in testis are significantly under-represented among genes linked to recessive Mendelian disorders (Fisher’s Exact Test p=1.0x10^-40^; Fig. 3d).

### Integrative analysis with testis scRNAseq reveals ‘molecular subforms’ of NOA

We next aimed to characterize the testicular cell types and pathways affected by prioritized NOA variants by characterizing scRNAseq data from twelve human testis samples (Methods) ^27^. Testis expressed genes were grouped into 70 expression modules (“components”) by soft clustering with Sparse Decomposition of Arrays (SDA), a Bayesian method for tensor decomposition (Fig. 4a, Methods). Each component is described by two matrices: a cell score matrix and a gene loadings matrix. Together they provide a quantitative summary of which genes are co-expressed across which cells. We provide an interactive browser to view the SDA components in detail online at https://conradlab.shinyapps.io/HISTA/#. These components represent sets of genes with similar expression dynamics independent of cell type boundaries and are often driven by shared biology ^28^. The activity of each component can be summarized by plotting the cell scores for each component on a map of labeled types (Fig. 4b), and, in the case of germ cell components, plotting cells scores as a function of developmental stage (pseudotime, Fig. 4c). In such a way components can be identified that are specifically expressed in spermatogonia (component 35), spermatocytes (component 59) and spermatids (component 92) (Fig. 4c).

**Fig. 4.**
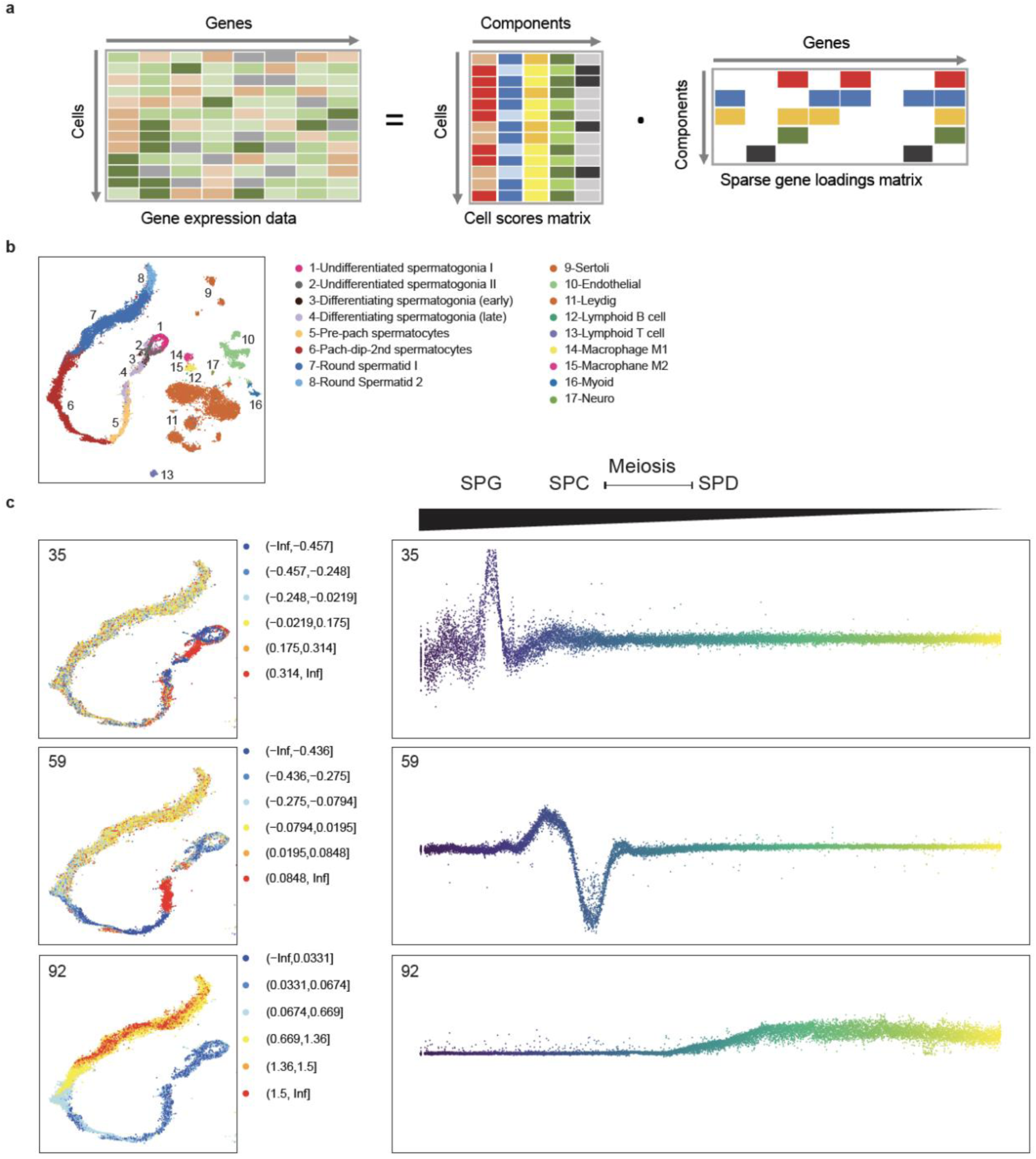
Decomposition of testis gene expression patterns with sparse decomposition of arrays (SDA). **a,** We applied sparse decomposition analysis (SDA) to testis scRNA-seq data from 12 human donors to identify latent factors (‘components’) representing gene modules. These components are defined by two vectors – one that indicates the loading of each cell on the component, and one that indicates the loading of each gene on the component. **b,** the same scRNA-seq data was summarized using a conventional tSNE analysis, and testicular cell types labels assigned to all cells. **c,** by plotting the cell scores for three representative germ cell components on the tSNE, and as a function of pseudotime, it is apparent that transcription during spermatogenesis can be modeled as series of components overlapping in time, coming on and off gradually on different timescales ^28^. Shown are components with activity that start in spermatogonia (35), spermatocytes (59), and spermatids (92).

By comparing the profile of SDA genes loadings for GEMINI NOA genes to the profile observed for 1) genes essential for spermatogenesis in mice based on MGI database and literature searches (denoted ‘MGI genes‘; n=197) and 2) genes identified among the controls of the institutional cohort (n=131), we observed that the mouse MGI genes were more similar to NOA rather than to the control genes (R=0.54 vs R=0.087; p=3x10^-4^ Pearson-Filon test, Fig. 5a). Unlike controls, NOA and MGI genes showed a very strong, specific enrichment on SDA components with peak expression in earlier phases of germ cell development, differentiating spermatogonia and pre-pachytene spermatocytes (Fig. 5a). Specifically, component 51 was an outlier in the absolute number of human and murine infertility genes (Fig. 5b-c) and showed strong enrichment of gene ontology (GO) categories related to meiosis. At least half of the top 50 genes in this component have already been identified as associated with male infertility in humans, mice, or both, whereas several of the genes with a high specificity towards this component have yet to be characterized in mammalian spermatogenesis, including *RAD51AP2*, *PRAP1*, *C18orf63*, *C5orf47*, and the *SYCP3* paralogs *FAM9B* and *FAM9C* (Fig. 5b).

**Fig 5.**
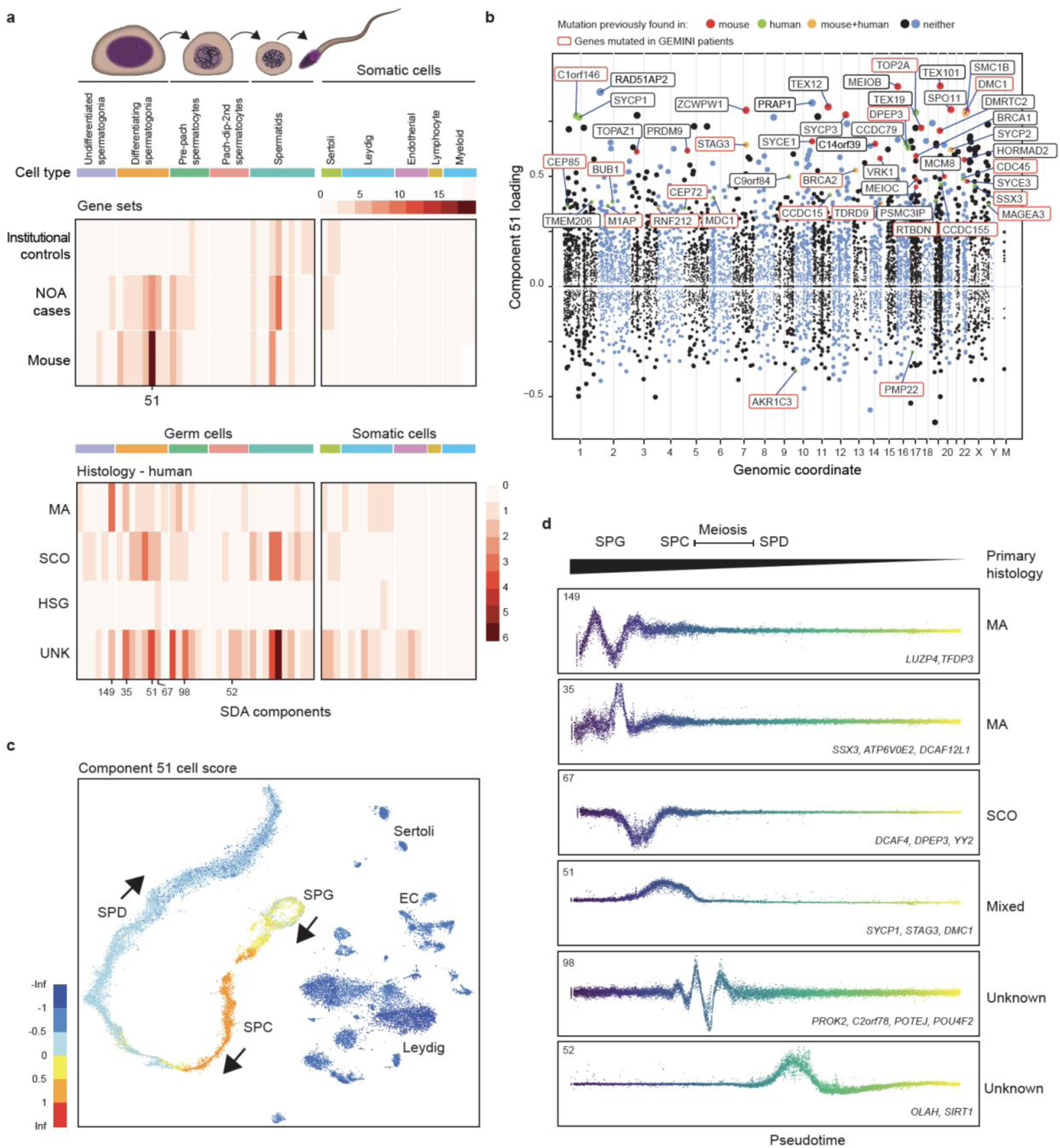
Using SDA components to define molecular subforms of genetic infertility. **a**, Heatmaps summarizing the count of genes loading on each SDA component. TOP: The distribution of NOA candidate genes across components is more similar to the distribution of mouse infertility genes, compared to genes with rare damaging genotypes in population controls (p=3x10^-4^, Pearson and Filon’s Z). BOTTOM: Distribution of NOA candidate genes by testicular histology across SDA components. Genes found in patients with MA show a different pattern of expression compared to genes in SCO patients. SPG, spermatogonia; SPC, spermatocytes, SPD, spermatids; UNK, undetermined histology. **b**, Expression of SDA component 51 visualized on a t-SNE representation of the human testis scRNA-seq dataset. This component is expressed in spermatogonia, and more highly in pre-leptotene spermatocytes. **c**, Gene loadings for SDA component 51. Gene loadings reflect which genes are active in a component, with stronger positive or negative loadings indicating greater expression changes in the component compared to the total dataset. **d**, Germ cell expression of components with multiple gene loadings ordered by pseudotime, shown with representative genes and patient histologies.

We next compared the SDA profiles in the context of the histological diagnosis of the NOA cases and observed a different distribution of gene loadings across the components for patients with MA compared to patients with SCO or “unknown” histology, the latter of which is likely a mixture of all three histologic subtypes (p=3.0x10^-3^, hypergeometric test, Fig. 5a). Furthermore, the human SDA profile for MA genes was negatively correlated with the profile of SCO-associated genes and genes from patients with unknown histology (R=-0.23) indicating distinct molecular causes for different histological subtypes. This observation of different loading patterns for genes linked to MA compared to SCO (p=2.8x10^-3^, hypergeometric test) was replicated with data from 197 mouse models of male infertility with known histological observations (Methods).

SDA components were classified into “MA-enriched” and “SCO-enriched” based on the corresponding case histology of component genes. Among the MA components with multiple gene loadings, SDA149 and SDA35 were expressed in undifferentiated and differentiating spermatogonia and involved multiple known early germ cell markers, such as *NANOS3, ID4* and *DMRT1,* but also *DCAF12L1,* a gene recurrently disrupted in NOA cases (Fig. 5d). The SDA94 component represents an alternative molecular time-point affected in MA cases, meiotic spermatocytes, where cases were identified with mutations in *HIST1H1T* and *C1orf146* (a human ortholog of yeast *SPO16*). Similarly, five SCO ‘molecular subforms’ were mapped to the spermatogonial cell population, including SDA67, enriched for DNA replication genes, and SDA46, a component with unknown function (see below). Collectively, these findings indicate that within the same histological endpoint, different ‘molecular subforms’ can be found reflecting the known and novel molecular origins of azoospermia.

### SDA components with uncharacterized genes

We noted several SDA components that included multiple prioritized genes not previously linked to mammalian sperm production. For instance, component 46, which included NOA case genes *TUBA3C*, *CDC45*, *FOXM1*, and *GTSE1*, cycles strongly two times in B spermatogonia, corresponding to the number of mitotic divisions these cells undergo in humans and which are thought to be the equivalent of transit amplification of stem cells in somatic tissues (Extended Data Fig. 7a-c)^29^. The number of mitotic divisions that occur during spermatogonial amplification appears to vary among species^30^, raising the fascinating possibility that there may be evolvable machinery that specifically controls germline, but not somatic, mitoses. Concordantly, we found that roughly half of the top loading genes on component 46 exhibited testis-enhanced expression (Extended Data Fig. 7d).

Component 98 included the prioritized genes *C2orf78*, *POTEJ*, *POU4F2*, and *PROK2*, and has a strong, specific expression pattern in spermatocytes (Fig 5d, Extended Data Fig. 7a-c). Loading in the top 60 genes of this component are 39 lincRNAs, and, in addition to *C2orf78*, three other uncharacterized protein-coding genes (*C17orf9*6, *C9orf163*, *C9orf57*). Three GO annotations are significant for this mysterious component, all indicating functions in plasma membrane cell-cell adhesion (GO:0007156, GO:0098742, GO:0016339, all p<10^-10^). Component 122 contains the NOA genes *INHBB*, *MAMLD1*, and *SMIM1*, the first two of which have previously characterized roles in gonadal function^31, 32^. This component is specifically expressed in Sertoli cells, raising the important possibility that some forms of human spermatogenic impairment may be ascribed to somatic cell defects (Extended Data Fig. 7a,c).

### piRNA processing factors in NOA

We prioritized potential disease variation in six genes that are essential for the biogenesis of PIWI-interacting RNAs (piRNA). piRNAs are small non-coding RNA molecules highly enriched within and critical for the survival of the germ cell pool through silencing transposable elements in fetal germ cells and for transcript storage and degradation in adult meiotic and haploid germ cells^33, 34^ (Fig. 6a). In male mice, disruption of any of the components of the piRNA biogenesis leads to detectable changes in the expression of mature piRNAs and causes spermatogenic arrest^35–41^. Concordantly, spermatogenic arrest was mostly characteristic to the eleven NOA cases who were affected by rare recessive variation in six piRNA biogenesis genes (*PLD6, PNLDC1, RNF17, TDRD9, TDRD12, TDRKH*; Extended Data Fig. 8a). *TDRD9* ^42^ and *RNF17* ^43^ have previously been found to be disrupted in men with azoospermia.

**Fig. 6.**
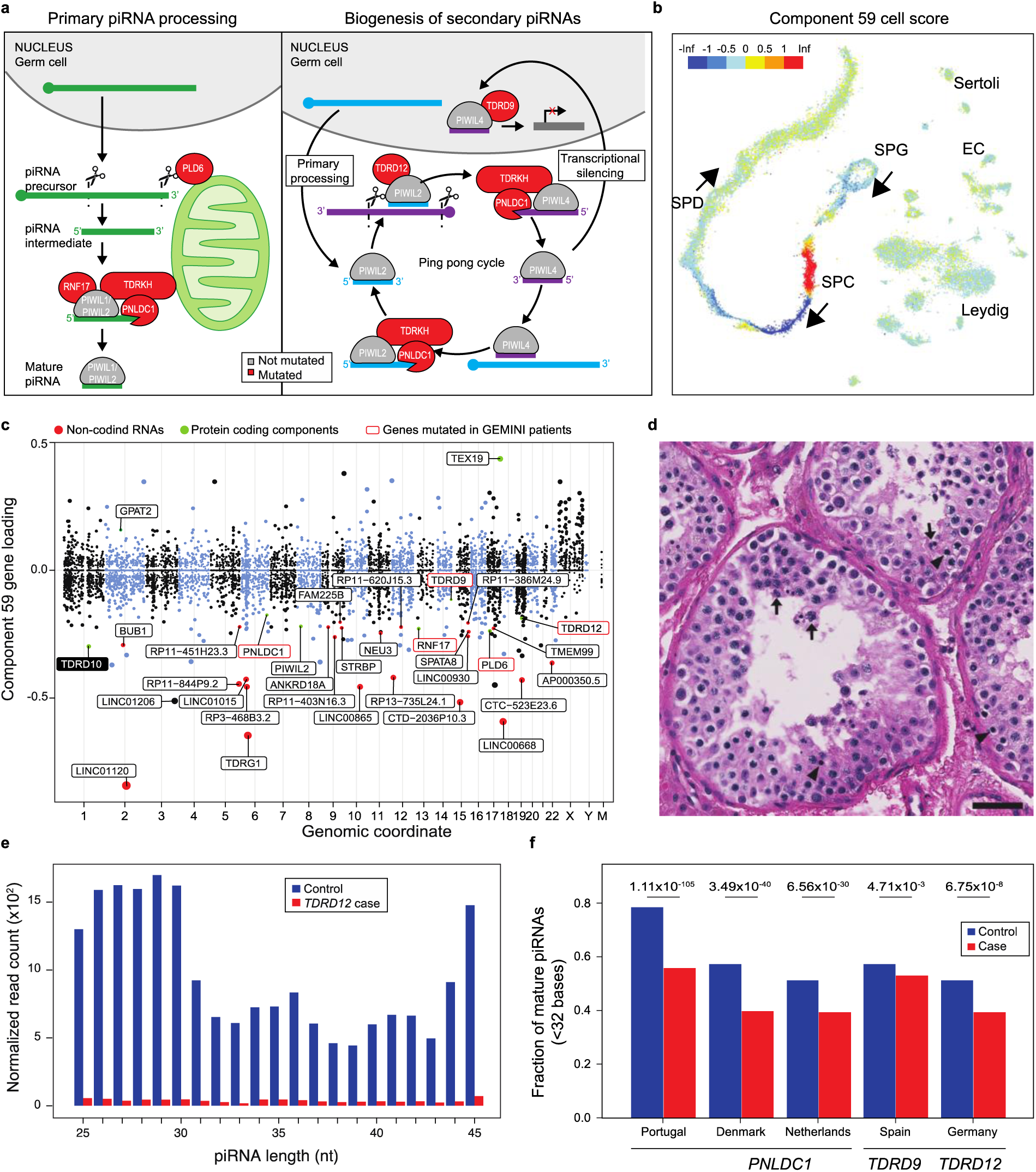
Disruption of piRNA biogenesis causes spermatogenic failure in men. **a,** A schematic of piRNA biogenesis with the components affected among NOA cases in this study highlighted in red. piRNAs are produced by two biogenesis pathways. The primary pathway involves transcription of long precursor-transcripts from genomic clusters, which are then processed in the cytoplasm. **b**, Cell scores for component 59, which encodes both piRNA processing genes and target pre-pre-piRNAs, indicate that it is expressed primarily in pre-leptotene spermatocytes. **c**, Gene loadings for component 59. Note *TDRD10* (black box), a protein-coding gene which has yet to be characterized for a role in mammalian piRNA processing. **d**, H&E stain of *TDRD9* patient biopsy showing spermatogenic arrest. The most mature germ cell observed were early round spermatids, which often appeared multinucleated (arrows). In addition, many pyknotic cells were observed (arrowheads). Scale bar = 50 *μ*m. **e**, Size distribution of piRNAs detected in the NOA case with a biallelic missense variant in *TDRD12* and matching control, derived from small RNA-sequencing of testis tissue. **f**, Significantly decreased fraction of mature (<32 bases) piRNAs was detected in testis of patients with piRNA pathway mutations, indicative of faulty processing of the immature piRNA transcripts.

Remarkably, when queried against the SDA components, these 6 genes loaded highly and specifically on component 59 expressed in pre-pachytene spermatocytes (p<0.01, Fig. 6b) together with other known piRNA processing genes *PIWIL2*, *TEX19* and *GPAT2*, and a potentially novel factor in piRNA processing, *TDRD10* (Fig. 6c). Strikingly, 11 of the 15 top loadings on component 59 were long intergenic non-coding RNAs annotated as pre-pre-piRNAs^44^, indicating that the precursors of piRNA molecules are strongly co-regulated with the transcripts of the piRNA biogenesis genes themselves.

We set out to validate the functional consequences in testicular tissue samples available for six NOA cases with prioritized variants in *PNLDC1, TDRD9* and *TDRD12*. *PNLDC1,* a trimmer of pre-piRNAs, was recurrently disrupted in three men from different GEMINI centers and one case from the Newcastle/Radboud cohort ^45^, whereas two potentially deleterious variants in *TDRD9* and *TDRD12,* essential factors in secondary piRNA biogenesis, were found in singleton NOA cases from the GEMINI and MERGE cohorts, respectively (Extended Data Fig. 8a). Disruption of all tested piRNA genes predominantly led to spermatogenic arrest seen in testicular biopsy (Fig. 6d, Extended Data Fig. 8a-c,e,f)^45^ and the altered piRNA biogenesis was clearly detectable upon small RNA sequencing with a shift towards longer immature piRNAs and significantly reduced fraction of mature piRNA molecules <32 nt compared to respective controls with normal spermatogenesis (1.1-1.3 fold decrease, p<0.01, hypergeometric test; Fig. 6e-f; Extended Data Fig. 8d,g; Supplementary Discussion) ^45^. Collectively, these findings demonstrate that components of piRNAs biogenesis are indispensable for human spermatogenesis and highlight the benefit of intersecting putative disease genes with scRNAseq data for identification of diverse molecular subforms of NOA.

## Discussion

In this study, we performed whole exome sequencing on an international cohort of unrelated NOA cases and reported extensive genetic heterogeneity attributable to the recessive form of the disease. We implicated over 200 genes in primary spermatogenic failure, including 27 genes with significant association with NOA when comparing 2,072 cases and 11,587 fertile controls. Thus, we provide two lists of genes with two levels of statistical signal: 221 genes prioritized in GEMINI cases, the majority of which were observed in n=1 cases, with an estimated 30% false discovery rate based on a control cohort comparison (Supplementary Table 2). Second, we provide a higher confidence list of 27 genes which have all been observed in multiple NOA cases and have statistically significant association with NOA in a comparison with 11,587 fertile controls (Supplementary Table 4). The observed detection rate of possible causes of NOA in GEMINI (20%) remains a conservative estimate, as our approach did not consider genes exclusively involved in fetal gonad development, excluded genes with broad expression across the body, and did not address dominant or additive defects likely to contribute to the manifestation of NOA as well ^46, 47^. A more detailed analysis of X-linked genes is reported in a companion paper focusing exclusively on the X chromosome, using a different method of prioritization than described here (Riera Escamilla et al. submitted).

One remarkable conclusion of our study is that the Mendelian causes of NOA are well distributed across the vast number of genes involved in testis function, and not clustered into a small number of genes. The highest level of recurrence recognized in the GEMINI cohort was n=3 cases affected by mutations in the same gene, and most cases were singletons, affecting in total 200 genes. This places NOA in a similar space of genetic architecture as congenital hearing loss (151 genes with at least “limited” clinical validity) ^48^, inherited retinal degenerations (over 260 genes) ^49^, and ciliopathies (over 190 genes) ^50^, but not yet approaching developmental disorders (over 1,500 reportable genes) ^51^. This observation has numerous implications for the clinical management of genetic forms of male infertility. First, it seems unlikely that a targeted panel of genes for clinical genetic testing will, in the short term, be an effective replacement for whole exome or genome assays in this disease space. Second, most clinicians will never see multiple unrelated cases of infertility caused by mutations in the same gene. Looking across clinics to share experiences for diagnosis and treatment, and to construct case series, will be essential to optimize patient care. From a research perspective, this scarcity of recurrent cases will challenge variant interpretation: this will need to be addressed by data sharing consortia, development of robust assays to evaluate the function of a wide range of variants, and more creative, integrative use of model organisms for functional validation.

It is now clear that what is clinically referred to as “spermatogenic impairment” is a collection of numerous rare diseases. However, these rare diseases share a related genetic basis; many of the affected genes encode proteins that either interact physically, such as in a protein complex, or physiologically, in regulating a particular cellular process. In such a system it is expected that oligogenic inheritance will occur, as well as variable expression of disease genotypes, where the precise phenotypic effect of a mutation will depend on genetic variation at other interacting loci, as well as the environment (e.g. patient age, lifestyle)^52^. We noted that eleven genes prioritized in our study have previously been linked to male infertility disorders other than NOA, including hypogonadotropic hypogonadism, primary ciliary dyskinesia, and multiple morphological abnormalities of the sperm flagella (MMAF; Fig. 3c, Extended Data Table 2, Supplementary Discussion) ^7, 53^. The latter suggests that problems in sperm assembly may also result in defects in sperm concentration, consistent with the observation of severe oligozoospermia in some patients with MMAF ^54, 55^; ^56^. Clinical data available for the affected GEMINI patients did not indicate misdiagnoses, and most of these genes are robustly expressed in adult testicular cell types relevant to NOA, raising the possibility of an expanded phenotype for these loci. In total, these observations suggest that variant and gene interpretation for NOA should typically be performed jointly with genetic knowledge of other male infertility phenotypes. It should be expected that, while some subtypes of NOA may have a unique genetic basis not shared with other traits, many variants that have a causal role in NOA will also modulate sperm production across the full spectrum of sperm count, such has already been shown with some Y chromosome variants ^57^. Furthermore, it is imperative that future studies of NOA should consider oligogenic inheritance during case assessment, and population-based tests of association that include heterozygous genotypes should be used as discovery and validation tools.

By intersecting the identified NOA disease genes with an scRNAseq atlas of human adult testis, we were able to organize the underlying molecular pathologies beyond what is currently possible with histological classification. We identified gene expression components with clear sets of independent genes, involving distinct cellular processes (e.g. mitosis, meiosis, piRNA biogenesis) and distinct cell types (e.g. Sertoli cells, and various germ cell types). In total we found over a dozen clearly distinct components with robust GEMINI gene loadings, and have statistically defined 70 components from total testis expression data. Thus the scope is clearly there to use gene expression components as a rational basis for NOA patient classification with higher resolution than our current systems based on histology. We are not proposing that gene expression signatures like SDA components will provide a comprehensive and definitive classification system for all genetic forms of male infertility, just that such an approach can provide a useful starting point for organizing the rapidly growing, and, in some cases, not obviously related genetic causes that are being identified.

The use of components to categorize patients can have benefits to research and patient management. Most importantly, we showed that defects in multiple genes from the same component, 59, produced a shared molecular signature of defective piRNA processing that could be detected with the same assay of small RNA sequencing. With further research, it may be possible to design similar companion diagnostics for each component, or molecular subform, that can be used in conjunction with genetic analysis to confirm the pathogenicity of candidate variants, even for variants and genes that have yet to be characterized. Examples of other molecular markers that could be measured from patient tissue would be DNA methylation (e.g. for *SPOCD1* mutations found in our study ^58^), metabolite levels, or markers of immune activity, although the potential number is large and many have been considered to date. The SDA approach simply provides a framework to pair genes of uncertain function with other genes, many of which may already have well characterized roles. Finally, we showed that components can be used to map the cell types involved in a defect. This would be critical knowledge to nominate patients with primary defects in somatic cells, who could benefit from powerful *in vivo* therapies targeted to those somatic cells, without directly modifying germ cells.

Infertility treatment is the leading frontier of some genetic technology, including diagnostic single cell sequencing ^59^ and therapeutic mitochondrial donation ^60^. The principles outlined in this discovery study indicate that genetic forms of male infertility have diverse mechanisms, and that genetic diagnosis through whole exome/genome sequencing will be an essential tool for developing and targeting new forms of assisted reproductive therapies but also for risk evaluation of future comorbidities.

## Subjects and methods

### Study subjects

Anonymized DNA samples were collected for a cohort of 1,011 unrelated men diagnosed with idiopathic NOA and recruited in four centers in the USA, two centers in Portugal (irreversibly anonymized) and a total of five centers in Australia, Canada, Denmark, Estonia and Spain. Thus, determination of NOA and infertility workup varied by practice pattern across centers, but in general followed published clinical guidelines of American Urological Association/American Society for Reproductive Medicine (AUA/ASRM) and European Academy of Andrology (EAA)^16, 17^. Men were confirmed to have azoospermia (no spermatozoa detected in the whole ejaculates) or severe oligozoospermia/cryptozoospermia (n=9 subjects; extremely low concentration of spermatozoa, <∼1 million/mL; jointly termed as ‘NOA’ in this study) according to the AUA/ASRM guidelines and based on physical examination (testis volume), endocrine measures (FSH, LH, and T) and histological findings if available. In the case of mixed findings on histology, i.e. the observation of MA tubules and SCO tubules in the same patient, we classified the patient as MA if any MA tubules were noted.

Testis volume and presence and grade of varicocele were evaluated by palpation or ultrasound by a trained fertility specialist. The exclusion criteria included the obstruction or absence of vas deferens, varicocele of bilateral grade 2-3 or unilateral grade 3, radical pelvic surgery, anejaculation, spinal cord injury, radiation treatments or chemotherapy exposure and environmental factors. CBAVD and other forms of obstructive azoospermia were ruled out based on a variety of factors including semen parameters (semen volume and qualitative fructose test), endocrine evaluation (normal FSH), normal testis volume and the presence of vas deferens detected by palpation and/or ultrasound. In addition, men with positive findings of Y chromosome microdeletions (YCMD) or karyotype abnormalities (including Klinefelter syndrome, 47, XXY) were excluded from the study. As a large fraction of the GEMINI cohort are historical cases, the information on the YCMD or 47, XXY testing was not available for all subjects. To retrospectively identify and remove cases where YCMD or 47, XXY karyotype was not originally assessed or was missed in the patient workup, we analyzed the genotype callset of the cases generated based on the WES data (Extended Data Fig. 1; further details in Supplementary Discussion). Structural abnormalities of sex chromosomes (including YCMD) and autosomes were detected from the copy number variation data, whereas cases of Klinefelter syndrome were identified by a combination of normalized coverage of chromosome Y and the inbreeding coefficient *F* of chromosome X (Extended Data Fig. 1c) and confirmed by the presence of large copy number variants on chromosome X.

To identify potentially missed cases of obstructive azoospermia, the genotype call set was additionally screened for pathogenic mono- or biallelic ClinVar variants (curated Expert Panel list, July 2018)^18^ in the *CFTR* gene, a known cause of obstructive azoospermia. We elected to remove cases who were heterozygous for a single well-acknowledged pathogenic *CFTR* variant as a conservative measure. Due to the limitations of WES to detect variants in non-exonic regions, we are not able to exclude the presence of other non-coding pathogenic variants in the *CFTR* gene in trans with the coding variants^19, 20^. Additionally, identity by descent (IBD) analysis was performed using *SNPRelate* R package^21^ to identify and remove one counterpart of each accidental sample duplicate or twin pairs and cases with cryptic relatedness to avoid ascertainment bias of rare variation.

### Institutional control cohort for development of variation prioritization procedures

A collection of individuals (total n=2265, 851 of whom are men) sequenced with the identical exome platform as the GEMINI cases was used as institutional controls to refine the variant prioritization pipeline and to test its ability to specifically aggregate potentially pathogenic variation related to testicular function. This comparative dataset consisted of 79 patients from Alzheimer study as well as 2186 participants from a study of skeletal or brain malformations: cases with adolescent idiopathic scoliosis (AIS, n=1245), Chiari malformation (n=105), hypermobility (n=38) and/or limb abnormalities (n=798), and 43 controls with no malformations. The AIS dataset has previously been partially reported^22^ and is fully available through the database of Genotypes and Phenotypes (dbGaP; https://www.ncbi.nlm.nih.gov/gap/; accession ID phs001677.v1.p1). The institutional cohort was subjected to exome capture and sequencing methods identical to the GEMINI cases (McDonnell Genome Institute, St. Louis, MO, USA) and similar bioinformatic pipeline was applied to read alignment and joint genotyping procedures. The prioritization of variants and genes likely disrupting testicular function was performed in parallel with the GEMINI cases and was aiming to refine the prioritization strategy in an inheritance mode-aware manner to maximize patient-specific calls. Genes with prioritized variants in both cohorts were removed from both as a conservative step.

### Additional cohorts

Two independent “replication” cohorts were screened for variation in genes linked to NOA in the GEMINI study. Whole exome sequencing of 900 men has been performed in the framework of the Male Reproductive Genomics (MERGE) study and includes cases with NOA or extremely low sperm counts (n=817). Patients were recruited at the Centre of Reproductive Medicine and Andrology (CeRA), Münster, Germany and the Clinic for Urology, Pediatric Urology and Andrology, Gießen, Germany. The MERGE cohort was queried for the 221 prioritized GEMINI genes to identify potential disease variation and retain recessive changes that are rare (MAF<0.01), non-synonymous variants seen in the MERGE cohort less than 20 times, had alternate read count <5, frequency >25 in all reads and CADD score >10 if the consequence is missense. The data set used to compare to the GEMINI study was further filtered to match the GEMINI filtering procedures by excluding variation with PSAP P-value >10^-3^, the positions that were observed in more than three individuals and the compound heterozygous pairs that were found within a 10 bp proximity or occurred in more than one individual.

Whole exome sequencing in the Newcastle/Radboud cohort was performed on a total of 331 patients who presented with idiopathic NOA (n=164) or severe to extreme oligozoospermia (with or without asthenozoospermia; n=167) at the Radboudumc outpatient clinic between July 2007 to October 2017 (n=286) and at the Newcastle Fertility Clinic between January 2018 to January 2020 (n=45). The presence of chromosomal anomalies, AZF deletions or pathogenic *CFTR* variants were exclusion criteria, all patients remained idiopathic following thorough clinical evaluation. Variants were filtered for recessive changes that were rare (MAF<0.01), non-synonymous, seen in less than 5 patients, with an alternate read count >5, and an alternate allele frequency >15%, predicted to be pathogenic by 3 or more of the following variant prediction tools: SIFT^23^, PolyPhen-2^24^, MutationTaster^25^, FATHMM^26^, Mutation Assessor^27^ and CADD^28^.

All variants with read depth less than 50 were validated by Sanger sequencing.

### Ethics statements & IRBs from centers

The study was approved by the Ethics Committee of all collaborative centers: protocols #201107177 and #201109261 approved by the institutional review board (IRB) of Washington University in St. Louis, USA; IRB_00063950 approved by IRB of University of Utah, USA; PTDC/SAU-GMG/101229/2008 approved by the Ethics Committee and Hospital Authority, University of Porto, Portugal; 16030459 and 0102004794 approved by the IRB of Weill Cornell Medical College, New York, USA; Ref. No.: 2014/04c approved by the IRB of Fundació Puigvert, Barcelona, Spain; NL50495.091.14 version 4 approved by the Research Ethics Committee of the Radboud University, Nijmegen, The Netherlands; Ref: 18/NE/0089, Bursa: 05.01.2015/04 approved by the University Research Ethics Committee of University of Newcastle, UK; and ethical approvals from human ethics committees of Monash Surgical Day Hospital, Monash Medical Centre and Monash University, Australia; the ethics committee for the Capital Copenhagen Region (Ref. Nr. H-2-2014-103) and the Danish Personal Data Protection Agency (Datatilsynet 2012-58-0004, local Nr. 30-1482, I-Suite 03696) gave ethical approval for this work in accordance to the European Commission Directive for the transfer of personal data (MTA/I-4728.A1); the Ethics Committee of National Institute of Health Dr Ricardo Jorge, Lisboa, Portugal gave ethical approval for this work; and approval 74/54 (last amendment 288/M-13) released by Research Ethics Committee of the University of Tartu, Estonia. The MERGE study protocol was given ethical approval by the Ethics Committee of the Ärztekammer Westfalen-Lippe and the University of Münster (Ref. No. 2010-578-f-S). Written informed consent was obtained from all men.

### Whole exome sequencing and analysis

Whole exome sequencing of the genomic DNA extracted from blood was performed at McDonnell Genome Institute of the Washington University in St. Louis, MO, USA (genome.wustl.edu) using an in-house exome targeting reagent capturing 39.1 Mb of exome and 2x150 bp paired-end sequencing on Illumina HiSeq 4000. A subset of cases (5%) were processed with either Nextera Rapid Capture (Illumina, San Diego, USA) or Nextera Rapid Capture Expanded Exome kit according to the manufacturer’s protocol and sequenced on HiSeq 2500 2x101 bp or HiSeq 3000 in 2x150 paired-end mode. On average, 99.9% of reads mapped to the target regions yielding an average exome coverage of 80x across sequenced individuals and platforms.

Raw sequencing reads were processed and aligned to GRCh38 assembly in an alternate contig aware manner using bwa-mem v0.7.17^29^, Genome Analysis Toolkit v3.6.0^30^ (GATK) and Picard tools v2.10.0 (http://broadinstitute.github.io/picard/). All sequenced cases were tested for sample contamination with verifyBamID v1.1.3^31^. Joint genotyping of all samples was performed with GATK tools and following the best practices of GATK. In order to achieve a high-quality call set, only samples with contamination FREEMIX <5%, average coverage >30x, low excess of chimeric reads (<5%) and call rate >85% were included. Positions with high exome capture kit-specific missingness (>=15%), low InbreedingCoeff (< ’-0.2’) and multi-allelic insertions and deletions (INDELs) were filtered out, as well as individual genotypes with DP<10, GQ<30 or heterozygous variation in non-PAR regions of chromosome Y. Precision and recall rate of detected variation was calculated in relation to the CEPH individual NA12878 run in parallel with the study cohort and for which a reference call set of high-quality variants is publicly available (Illumina Platinum Genomes^32^). The genotyping achieved precision of 99% for SNVs and 91% for INDELs and a recall rate of 89% and 47%, respectively.

### Detection of copy number variation

Copy number variants were detected from WES data using XHMM as previously described^33^. CNVs were called using the Viterbi HMM algorithm with default XHMM parameters, and XHMM CNV quality scores were calculated as previously described using the forward– backward HMM algorithm. In addition, all called CNVs were statistically genotyped across all samples using the same XHMM quality scores and output as a single uniformly called VCF file.

The output of CNV calls was first separated into deletions (DEL) and duplications (DUP) and each subset was then filtered by frequency to remove common CNVs, which were present in >1% of individuals and defined as overlapping more than 50% of their respective targets. Only CNVs with quality scores greater than or equal to 60 were included in the downstream analysis. As previously recommended^34^, individuals having a CNV count greater than 3 SD above the mean (27 DELs or 30 DUPs, in this study) were removed from the analysis.

To evaluate the sensitivity of CNV detection from the WES dataset, a comparative CNV call set for the reference sample NA12878 sequenced in parallel with the GEMINI study subjects, was available through the whole genome sequencing (WGS) effort of the 1000 Genomes phase 3 project^35^. The CNV calls of the 1000G dataset were required to overlap with WES CNVs by at least 50% of length and span the exonic regions targeted in this study. The sensitivity estimate reached 13% (n=11/85 of reference CNVs), which is within the expected range considering the fragmented coverage of the genome in WES approach and comparable to previously reported sensitivity estimates (7.67%) for CNV calling from exome sequencing data with XHMM when compared to WGS^36^. In total, approximately 55% of the autosomal duplications (n=5/9) and 60% of autosomal deletions (14/23) identified in NA12878 in this study are supported by previous studies^35^ and are likely true positives.

### Validation of CNVs by PCR

A subset of likely homozygous/hemizygous deletions was selected for experimental validation by +/- PCR, performed with at least one primer pair located between the predicted deletion breakpoints (internal) and a second primer pair outside, either upstream or downstream from the deletion breakpoints in a region unaffected by the predicted deletion (external, positive control). CNV was confirmed when the internal set of primers did not result in amplification and the external set of primers outside the deletion successfully amplified from patient DNA. Both sets were required to amplify from control DNA. Polymerase reaction for +/- PCR was performed using 50ng of DNA and Qiagen HotStarTaq (Qiagen, Hilden, Germany). Deletions were considered as true positives when only amplification for the external primer pair(s) was obtained. The primer sequences and annealing temperatures are available upon request.

### Fine-mapping of the *STRA8* deletion

A homozygous ∼6kb deletion on chromosome 7 (chr7:134,925,271-134,931,454, hg19) residing within the *STRA8* gene was identified in an NOA case GEMINI-295 by calling CNVs from WES data. We first validated the presence of this deletion in the patient DNA using +/- PCR with primers anchoring inside the deletion in exons 2-4 as well as outside the deletion in exon 8. Only the second set of primers outside the deletion amplified from the patient DNA as expected (Fig. S6). We then tested four additional pairs of primers in the intronic regions flanking the deletion to further delimit the breakpoint. Finally, we designed primers for amplification across the expected breakpoint (gap-PCR) with a fragment of maximum expected size of 13 kb without the deletion. This primer set only resulted in a single product of ∼5 kb in the patient indicative of a homozygous deletion (Extended Data Fig. 5). The acquired PCR product was sequenced and allowed us to determine the precise localization of the deletion breakpoints at chr7:134925172-134933449 (hg19). The 5’ breakpoint is nearly equidistant (∼400 bp) from an L1ME3 repeat of the L1 family and an MLT1AD repeat of the ERVL-MaLR family.

PCR was performed using 50ng of DNA and Qiagen HotStarTaq (Qiagen, Hilden, Germany). The long PCR spanning the deletion breakpoint was performed with NZYLong DNA polymerase (Nyztech, Lisbon, Portugal) at 62°C and 6 minutes of annealing (35 cycles). The ∼5 kb PCR product was cloned into TOPO-TA vector (MilliporeSigma, St. Louis, MO, USA) and a clean sequence was obtained, which confirmed the breakpoints. Finally, we attempted to find heterozygous positions in the regions adjacent to the breakpoints to allow allele-specific analysis and confirm the existence of two deletions with exactly the same breakpoints but we did not find any heterozygous positions in either of the introns sequenced (1, 7 and 8). Primers are provided upon request.

### Variant prioritization

To prioritize likely pathogenic SNVs, INDELs and CNVs detected from the WES data, a modified version of the Population Sampling Probability (PSAP) software was utilized, which identifies genotypes that are unusual in the context of known human variation (https://github.com/conradlab/PSAP/)^37^. PSAP evaluates the probability of sampling a genotype or a set of genotypes based on the pathogenicity scores and frequencies of variants observed in the unaffected population. Unlike classical workflows of case-control studies, PSAP enables the identification of potential causal variants from a single genome without the need for matching control samples. PSAP takes into account the sex and the ethnicity (European, African and ‘other’) of the cases and tests for autosomal Mendelian disease models (autosomal dominant, single-variant autosomal recessive and compound-heterozygote autosomal recessive) but also X- and Y-linked inheritance patterns. The impact of nucleotide changes positioned within multiple overlapping genes is assessed independently for each gene. The genomic positions of detected variation were lifted over to human genome assembly hg19 supported by PSAP.

The prioritized variants were subsequently filtered to maximize true positive calls, very pathogenic rare changes and genes most likely to impair spermatogenesis. Variation was excluded if it had PSAP P-values >10^-3^, minor allele frequency >0.01 across all populations in the gnomAD database v2.1.1 (popmax, https://gnomad.broadinstitute.org/) or was common in the study cohort. Additionally, genes enriched for rare pathogenic variation (PSAP P<10^-4^, popmax MAF<0.01) and identical homozygous or hemizygous recessive changes seen in >3 individuals were removed. Compound heterozygous variants were subject to exclusion if identical pair of genotypes was observed in more than one case or were found in cis configuration as determined by pHASER v1.1.1^38^, in-house parsing of variant annotation in the VCF file or manual check of read alignments using IGV tools^39^.

In order to further aggregate variation most likely increasing Mendelian disease risk and affecting spermatogenesis pathways, the following information was considered: 1) loss-of-function (LoF; nonsense, splice site and frameshift variants) changes, 2) genes known to cause male infertility in mice (Mouse Genome Informatics, MGI, http://www.informatics.jax.org/) or implicated in male infertility in humans (at least ‘limited’ supporting evidence, n=164)^7^, and 3) a list of genes with elevated expression in testis retrieved from the Human Protein Atlas v18.1^40^ (n=2237). This dataset is a compilation of 172 individual samples, corresponding to 37 different human tissues analyzed by bulk RNAseq^41^. The Human Protein Atlas project compared the gene expression of ten normal testis samples to 162 other tissue samples (36 tissue types) to identify the list of genes with elevated expression in testis. The genes were further grouped based on the level of tissue specificity: tissue enriched (expression in one tissue at least 5-fold higher than all other tissues), group enriched (5-fold higher average TPM in a group of two to seven tissues compared to all other tissues) and tissue enhanced (5-fold higher average TPM in one or more tissues compared to the mean TPM of all tissues)^41^. All three groups were considered for prioritization of genes with a likely impact on testicular function.

The criteria for aggregating most plausible NOA variation were evaluated individually for each Mendelian disease model in parallel with the cohort of institutional controls aiming to maximize patient-specific calls. All autosomal variants were required to meet at least one of the three aggregation criteria mentioned above. X-linked variants were expected to be located in genes with testis enhanced expression and additionally meet either the prioritization criteria 1) or 2) mentioned above, whereas Y-linked variants were included if defined as LoF. Any NOA gene, which did not display testis enhanced expression based on the Human Protein Atlas dataset, was required to at least show an appreciable expression specifically in premeiotic germ cells or somatic cells of the testis (normalized TPM>0.5 in the testis scRNA-seq)^42^.

### Burden testing

We performed gene-based burden testing using the combined data from GEMINI, MERGE, and Newcastle/Radboud case cohorts, compared to a control cohort of 11,587 fertile parents sequenced at Radboud University. These fertile parents were ascertained from the genetic diagnostic lab at Radboud, where they were referred for genetic testing as part of clinical care for their children. The cohort consisted of approximately equal numbers of men (5,803) and women (5,784), with an ethnic makeup reflecting the local Dutch population. Due to data use limitations, it was not possible to analyze compound heterozygous variants in the control cohort; therefore, burden testing was only performed using single-variant genotype calls. Testing was performed on the 35 genes with more than one prioritized homozygous (autosomal) or hemizygous (X and Y chromosomes) variant in cases (i.e. a smaller number of recurrent genes than observed in the full data). Case and fertile control genotypes were processed using the same filtering and prioritization pipeline described above, so the only variants included in the analysis were the short list of prioritized thought to represent potential Mendelian causes of NOA. The proportion of individuals with filtered variants was compared between cases and controls using the Fisher Exact test, and q-values were obtained using the Benjamini-Hochberg method. Two forms of burden test were performed and reported - one test using just male control samples, and one using the combined male and female control cohort.

### Bioinformatic annotation

To determine the ancestry of GEMINI cases, EthSeq software^43^ was implemented using Phase 3 genotype and population data from the 1000 Genomes Project as a reference^35^. Only variants with MAF>0.2 and located within the exonic regions targeted in the GEMINI WES study were considered. Long runs of homozygosity (ROH) were detected using the H3M2 tool specifically designed for analysing WES data^44^. ROH detection was performed separately for European, African, East Asian and South Asian populations with default H3M2 parameters (DNorm=100.000, p1=0.1, p2=0.1, F=5). To identify the longest class of autozygous stretches most likely related to disease and reflecting recent inbreeding^45^, unsupervised five-component clustering was performed in a population-specific manner using *Mclust* (*mclust* package v.5.4.3 in R). Only the longest class (class 5) of the ROH regions reflecting recent consanguinity and most likely contributing to disease^45^ were considered when calculating the fraction of the autosome being homozygous (FROH). The long ROH regions were intersected with CNV calls to exclude hemizygous regions.

To extract high quality LoF calls predicted to cause gene knock-outs, homozygous, hemizygous and compound heterozygous LoF variation were annotated and filtered using the LOFTEE tool (v1.0.3; https://github.com/konradjk/loftee) implemented in Variant Effect Predictor (v99)^46^ as per Karczewski et al. 2019^47^. Comparative population frequency of human “knock-outs” for the respective genes was acquired by screening large variation databases for LoF variation and performing LOFTEE filtering if not already previously applied internally by the databases. The screened databases included gnomAD (v2.1.1. and v3; accessed 02/14/2020), deCODE (Supplemental table S4 in Sulem et al. 2015)^48^ and Human Gene Mutation Database (HGMD; 12/20/2014 data freeze)^49^. Additionally, Born in Bradford, Birmingham project, and East London Genes and Health (http://www.genesandhealth.org/)^50^ exome data was aggregated.

The functional protein association network with the prioritized list of disease genes was built using protein-protein interaction data of the STRING database v11.0 (stringApp v1.5.0) integrated in the Cytoscape platform (v3.7.2)^51^. Only high confidence interactions (>0.7) were considered. Genes located on chromosome Y are not mapped by the STRING database and are not included into the analysis. The resulting network was visualized using a prefuse force directed layout based on the protein-protein interaction scores.

The median gene expression (TPM) for genes of interest across 52 human tissues, including testis, was extracted from the The Genotype-Tissue Expression (GTEx) V8 RNA-Seq dataset^52^. To explore the expression patterns of GEMINI disease genes across GTEx tissues in comparison to other recessive Mendelian disorders, a list of genes (n=2588) linked to Mendelian diseases was retrieved from the Centers for Mendelian Genomics dataset (http://mendelian.org/phenotypes-genes; accessed Oct 01, 2020). The gene list was then intersected with an OMIM dataset (retrieved May 15, 2020) to only extract genes that follow a recessive inheritance pattern, which includes autosomal as well as X-linked and Y-linked recessive modes. GTEx expression values were available for 604 genes out of the 615 defined as recessive Mendelian disease genes and the difference in the expression levels, when compared to the GEMINI disease genes, was evaluated using Mann-Whitney U test (p-value below 0.05 was considered significant).

To explore the functional role of prioritized disease genes in mice, information on the reproductive phenotypes of knock-out mouse models was extracted from the MGI database, International Mouse Phenotyping Consortium (www.mousephenotype.org)^53^ and literature.

To estimate *p,* the number of monogenic azoospermia genes in non-consanguineous men, we used monte carlo simulations to evaluate a model that relates *p* to the expected rate of recurrent gene hits in a sequencing study, which we denote as *r.* We assume that for a given set of azoospermia genes, each gene in the set is equally likely to be a monogenic cause of disease (e.g. the amount and effect size of disease variation in each gene is the same). Next, we evaluate the likelihood of a particular value of ***p*** using the difference in the expected rate of recurrent hits, r, and the observed rate r_o_ (9.4% among non-consanguineous NOA cases) as an objective function. We searched over a grid of 1000 values of *p_i_*, i ranging from 1 to 1000, with *p_1_* = 1, *p_2_*= 2, and so forth. For each value of *p_i_*, we simulate a set of “gene hits” by random sampling with replacement 221 genes, the actual number of prioritized genes observed in GEMINI, from the pool of *p_i_* genes. This is done 1000 times for each value of *p_i_*, each time recording the corresponding value of *r_i_*, and then averaging across all 1000 replicates to arrive at a final estimate for r_i_. The objective function, *r_i_*-*r_O_*, was minimized at i=625. The sensitivity of detecting multiple gene hits as a function of the cohort size was then calculated considering the estimated size of the azoospermia gene pool (n=625), discovery rates of 15% for outbred samples, 76% for consanguineous samples and 20% for the GEMINI cohort overall (8% of consanguineous cases) and assuming the absence of natural selection.

### scRNAseq analyses

We created an aggregated dataset of human testis scRNA-seq from multiple sources, comprising cells from a total of 12 human donors. These consisted of 7 adults^42^, 3 adults^54^ (GEO accession #GSE109037), and two juvenile samples^55^ (GEO accession #GSE120506). As described elsewhere, these datasets were integrated and analyzed using the sparse decomposition of arrays (SDA) framework^56^. SDA soft clusters genes and cells into latent “components”, which are then manually interpreted and separated into “technical noise” and “signal”. Components corresponding to technical noise are removed from the data, thus providing batch correction. For the analyses described, here, SDA was run with 150 components, half of which were removed as technical noise. Gene ontology enrichment analysis was performed on the top 250 genes from each component (from each side) using the enrichGO function from the clusterProfiler R package in which p-values are calculated based on the hypergeometric distribution and corrected for testing of multiple biological process GO terms using the Benjamini-Hochberg procedure.

### Small RNA sequencing and analysis

RNA was extracted from the fixed testicular tissue of the NOA cases with prioritized variants in genes involved in piRNA biogenesis and controls with complete spermatogenesis matched to each case by fixative. Small RNA sequencing libraries were prepared using CATS Small RNA-seq kit (Diagenode, Cat. #C05010040) and sequenced on the MiSeq platform (Illumina, San Diego, CA). Raw sequencing reads were processed according to the CATS Small RNA-seq protocol and reads with 25-45 bases in length were mapped to the human assembly hg19 using bowtie v1.0.1^57^ allowing one mismatch. The mapped reads were filtered to remove all reads mapping to small non-coding RNAs other than piRNAs (DASHR v2.0^58^) and the remaining reads were intersected with 205 piRNA loci previously identified in the adult human testis^59^. Individual piRNA counts were normalized to the spike-in cel-miR-39-3p (5’-UCACCGGGUGUAAAUCAGCUUG-3’) added to the samples in the first step of the sequencing library preparation. Spike-in reads were mapped to the human reference using bowtie v1.0.1 and allowing no mismatches.

### Sanger sequencing

The regions of interest were amplified using Hot Start Taq 2x Master Mix (New England Biolabs, Ipswich, MA, USA), 0.4 µM primers and a touch-down PCR program. The amplified PCR products were purified with MinElute PCR Purification kit (Qiagen, Hilden, Germany) or GFX PCR DNA and Gel Band Purification Kit (GE Healthcare, Marlborough, MA) and sequenced at Integrated DNA Technologies (Coralville, IA, USA). The primer sequences are available upon request.

### Immunohistochemistry

Testicular tissue was fixed in Bouin’s fixative, processed and stained with an antibody directed against STRA8 (Abcam ab49405, diluted 1:250) using the ImmPRESS detection kit (Vector laboratories, CA, USA) as described before^60^. Antigen retrieval was performed in a microwave in TEG buffer and the staining developed 20 min with AEC.

## Data Availability

Sequencing data that support the findings of this study are either in process of being deposited in dbGAP database and the accession numbers will be available by the time of publication.

## Acknowledgements

We thank all of the study participants and the numerous medical staff that enabled this study. We also acknowledge the contribution of the Lisbon clinical team - Carlos Calhaz-Jorge, Ana Aguiar, Joaquim Nunes, and Sandra Sousa (Unidade de Medicina da Reproducão, Hospital de Santa Maria, Centro Hospitalar Lisboa Norte, Lisboa, Portugal), Maria Graça Pinto and Sónia Correia (Centro de Medicina Reprodutiva), for patients’ clinical evaluation.

## Author contributions

L.N., A.M.L., K.I.A. and D.F.C. designed the study. L.N., A.M.L. and D.F.C. performed computational and statistical analysis. W.-L.C., B.M., T.L., E.M., L.K. and E.F. contributed to computational analysis. L.N., R.S., I.G., A.S., L.K. and K.A. performed validation experiments. W.-L.C. and C.A.G. provided and analyzed whole exome data from institutional controls. C.F., M.O., M.J.X., A.P., S..K., J.A.V. and F.T. participated in the replication studies. L.N., A.M.L. and D.F.C wrote the manuscript. L.N., A.M.L., K.V.-C., K.A. and D.F.C. generated manuscript figures. S.S., F.C., S.F., A.B., J.G., R.M., K.R.M., E.S.J., N.J., E.R.-D.M., K.A.J., M.P., J.I.S., M.L.E., T.G.J., J.M.H., D.T.C., P.N.S., M.O., M.L., C.K., A.R.-E., S.K., K.A. and K.I.A. provided patient samples and data. K.V.-C., N.J., E.R.-D.M., K.A.J., M.P., J.I.S., M.L.E., T.G.J., J.M.H., D.T.C., P.N.S., M.O., M.L. and C.K. contributed to the study design, discussion and manuscript preparation as the GEMINI consortium members. All authors reviewed the manuscript.

## Funding

National Institutes of Health of the United States of America grant R01HD078641 (D.F.C, K.I.A)

National Institutes of Health of the United States of America grant P50HD096723 (D.F.C.) National Health and Medical Research Council of Australia grant APP1120356 (M.O., D.C., K.I.A., R.M. and J.A.V.)

Spanish Ministry of Health Instituto Carlos III-FIS grant FIS/FEDER-PI20/01562 (CK, A.R.-E.). Estonian Research Council grants IUT34-12 and PRG1021 (M.L., M.P.)

ReproUnion and the Innovation Fund Denmark grant 14-2013-4 (K.A.)

German Research Foundation Clinical Research Unit ‘Male Germ Cells’ grant DFG CRU326 (C.F., F.T.)

The Netherlands Organization for Scientific Research VICI grant 918-15-667 (J.A.V.) Investigator Award in Science from the Wellcome Trust grant 209451 (J.A.V.).

FCT/MCTES, through national funds attributed to Centre for Toxicogenomics and Human Health—ToxOmics, grant UID/BIM/00009/2016 (J.G.)

National Institute of Mental Health of the National Institutes of Health grant T32-MH014677 (W.-L.C)

## Competing Interests

Authors declare that they have no competing interests.

## Additional Information

**Supplementary Information** is available for this paper.

Correspondence and requests for materials should be addressed to Donald F. Conrad.

Reprints and permissions information is available at www.nature.com/reprints

---

**Extended Data Fig. 1.**
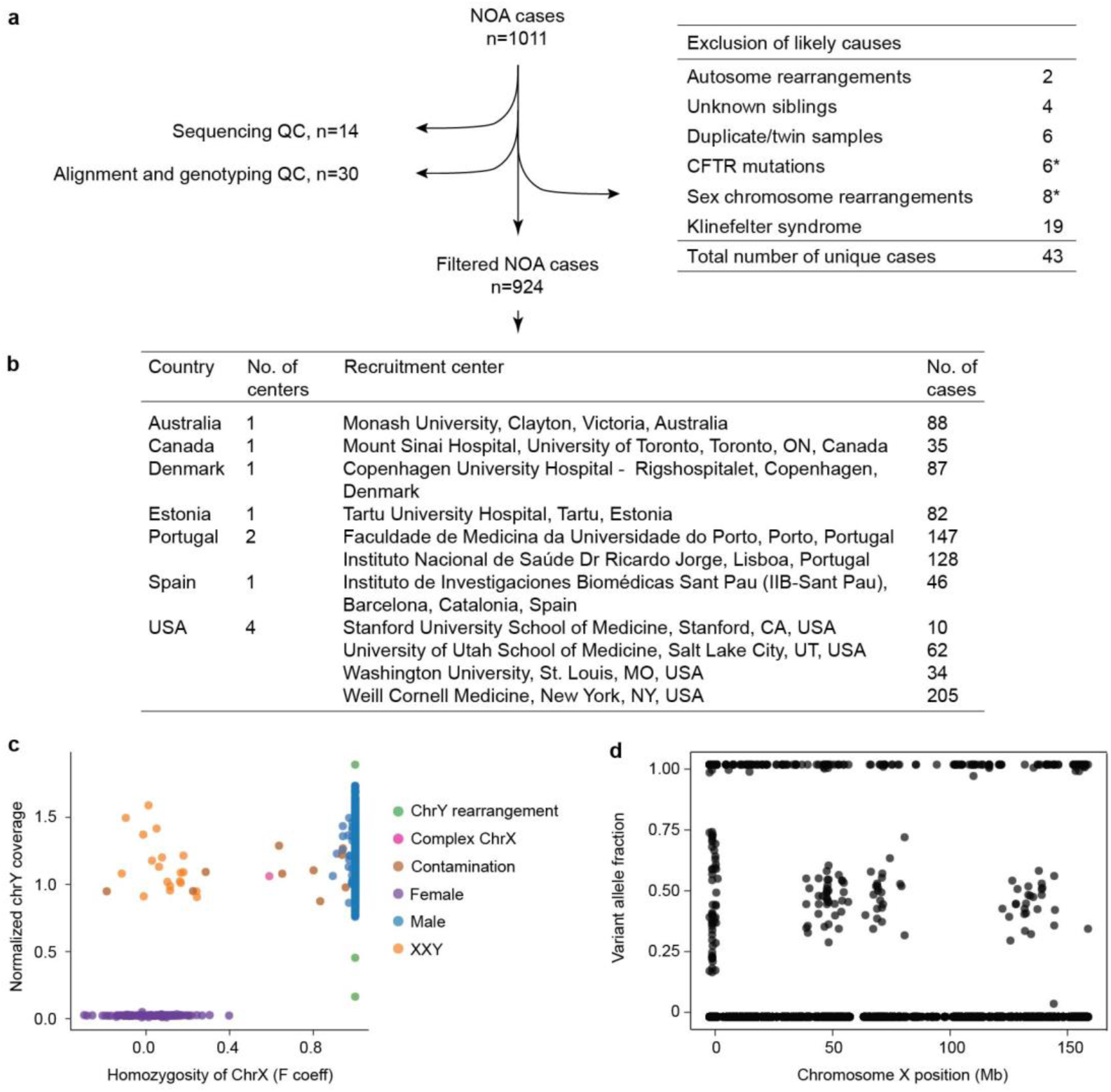
Filtering procedures of the GEMINI study cohort and screening for known genetic causes based on WES genotype call data. a, Sample filtering steps based on the WES data. Two cases harbored a combination of sex chromosome aberrations and a deleterious *CFTR* variant (asterisk; see Supplementary Discussion). b, Distribution of studied NOA cases (n=924) across the participating centers of the GEMINI study. c, Klinefelter syndrome (47,XXY karyotype) and large rearrangements on sex chromosomes were detected based on i) the CNV data and ii) the ratio of chromosome Y coverage (normalized to autosomes) and inbreeding coefficient *F* of chromosome X. Klinefelter syndrome was called if chrX *F*<0.5 and chrY coverage >0.6 and compared to the clustering observed for normal karyotypes of NOA cases (chrX *F*>0.5 and chrY coverage >0.6) and for women analyzed on an identical sequencing platform (n=96; chrX *F*<0.5 and chrY coverage <0.6). A NOA case with suspected large X chromosome rearrangements (‘Complex ChrX’) was identified as an outlier with respect to the inbreeding coefficient. Samples affected by DNA contamination (confirmed by FREEMIX tool, Methods) were not considered as candidates for sex chromosome aberrations. **d**, Distribution of variant allele fraction of all variants on the X-chromosome in the carrier of complex rearrangements.

**Extended Data Fig. 2.**
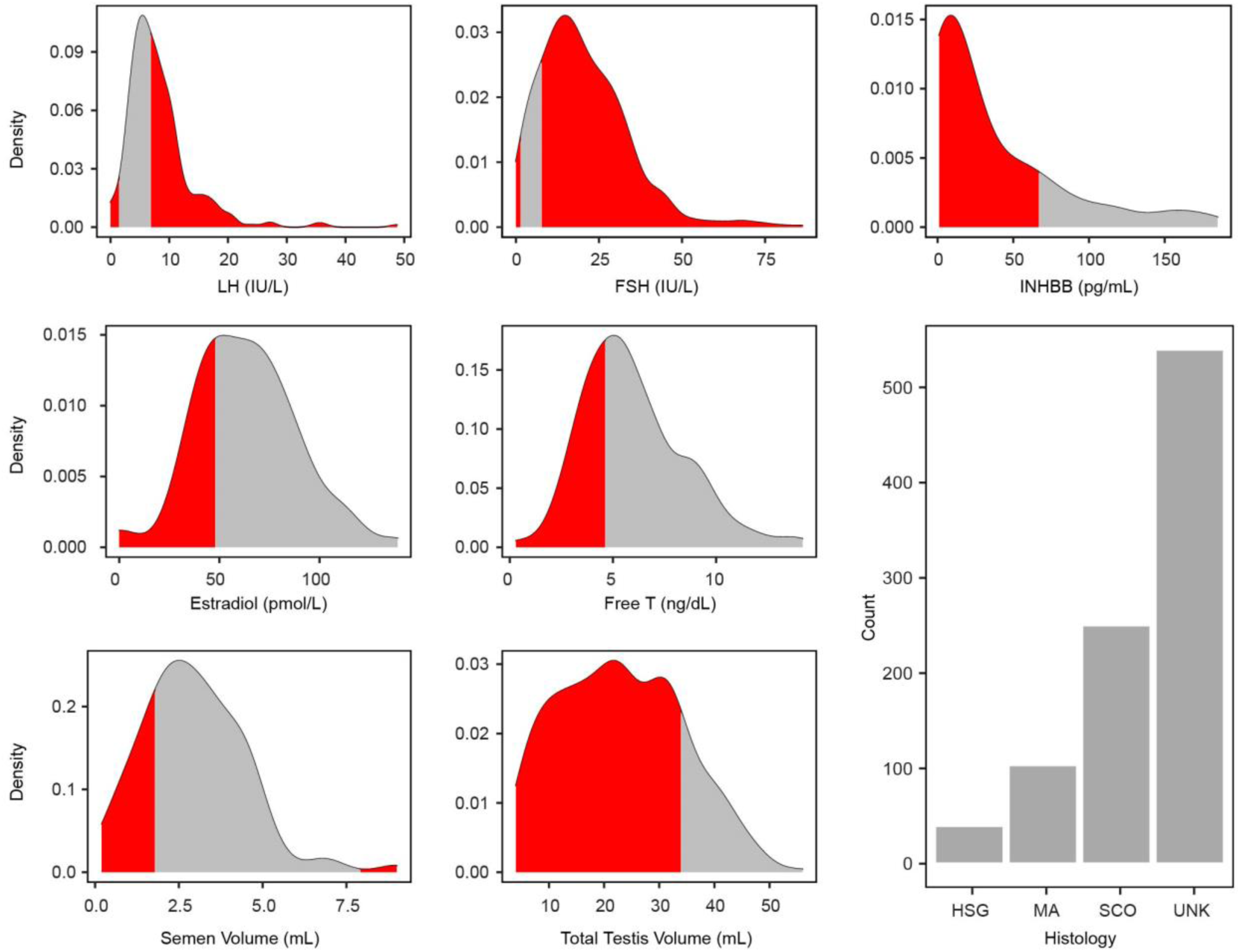
The distributions of reproductive hormone measurements, ejaculate volume and total testis size of 924 men included in the study. Measurements that fall outside of the published normal ranges (Supplementary Table 1) are shaded in red. HSG, hypospermatogenesis; MA, maturation arrest; SCO, Sertoli cell only; UNK, unknown testicular phenotype.

**Extended Data Fig. 3.**
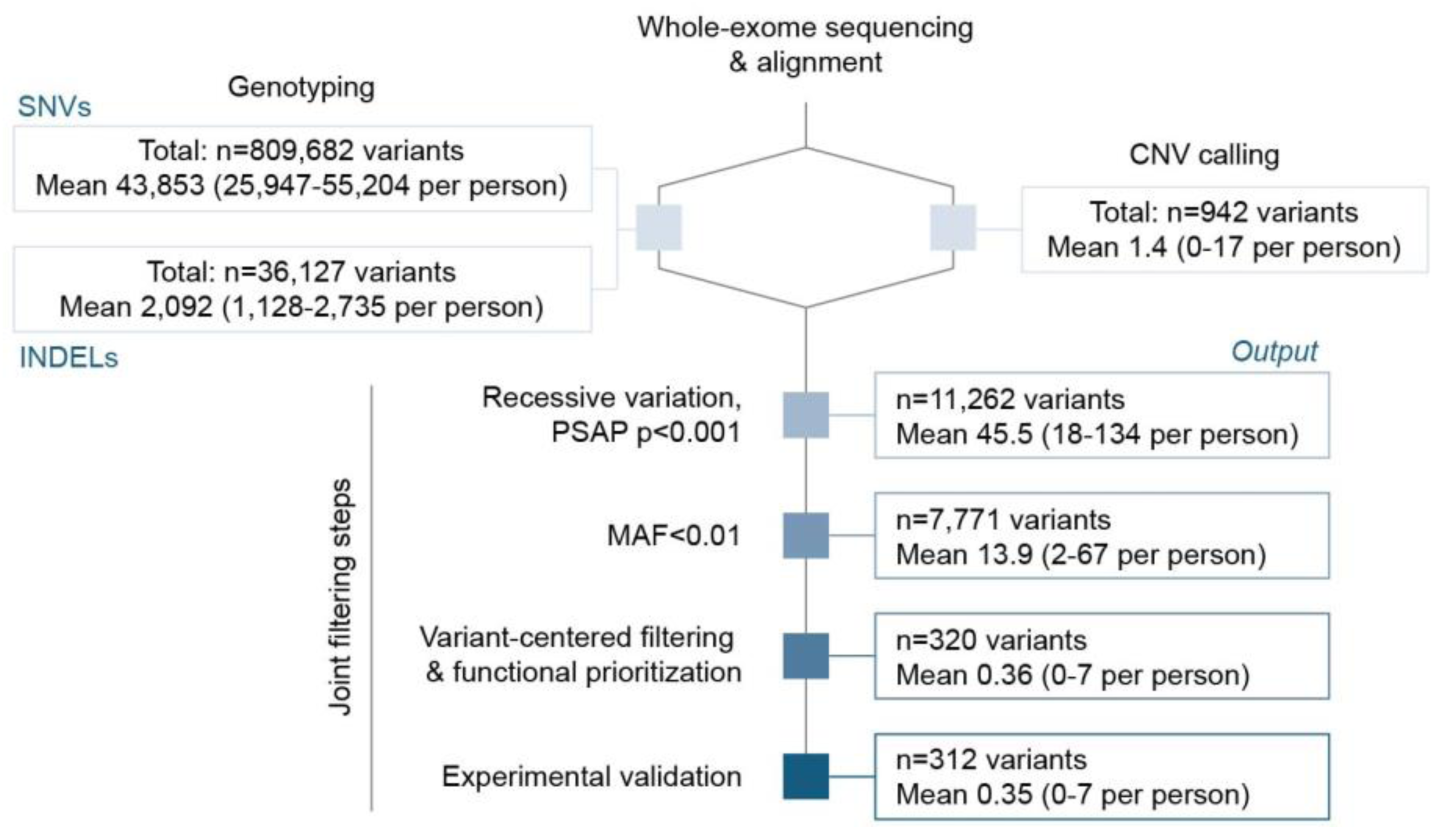
Detection and filtering of variants in 924 NOA cases included in the study. Whole-exome sequencing alignment was followed by detection and quality filtering of SNVs/INDELs and CNVs independently and subsequently combined for downstream filtering steps. PSAP was used to prioritize likely pathogenic genotypes, followed by selection of rare variation and exclusion of likely false positives (see details in Methods). The functional prioritization aimed to aggregate variation most likely to disrupt spermatogenesis. Total of 40% of all prioritized genotypes, predominantly LoFs and recurrent or poorly covered positions (read depth<20), were subjected to experimental validation by Sanger sequencing (SNVs and INDELs) or +/- PCR amplification (CNVs; Methods).

**Extended Data Fig. 4.**
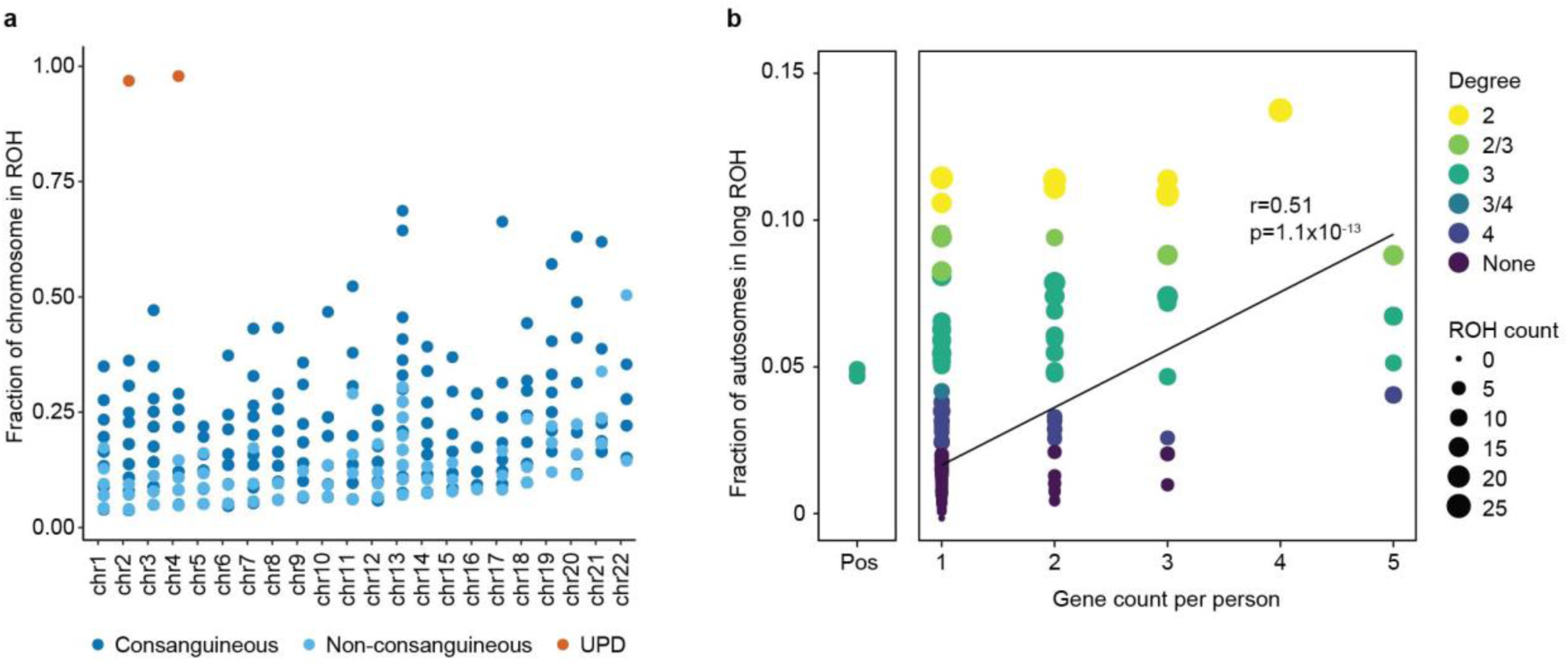
Uniparental isodisomy (UPD) and consanguinity in NOA cases. **a**, The fraction of chromosomes found in long runs of homozygosity (ROH) highlight two cases with UPD of chromosomes 2 and 4. **b**, Multiple disease genes are more likely to be identified in patients with multiple long runs of homozygosity (ROH) (r=0.51, P=1.1x10^-13^). Degree, estimated degree of parental relationship; Pos, siblings with known 3rd degree of consanguinity.

**Extended Data Fig. 5.**
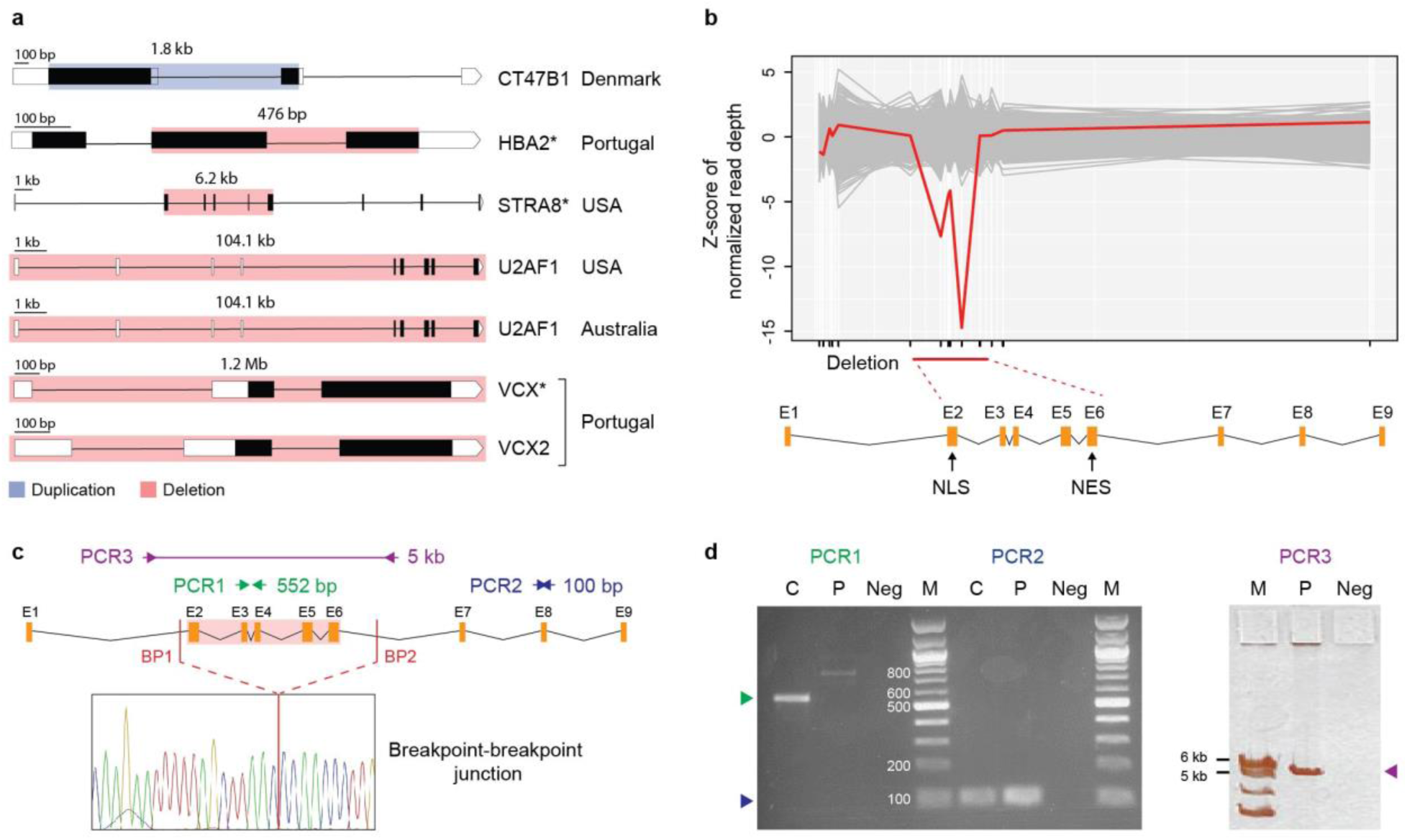
Prioritized copy number variants in NOA cases and validation of *STRA8* deletion. **a**, CNV events involving prioritized NOA genes. *HBA2* gene is exclusively expressed in the whole blood (GTEx database) and likely represents a false positive finding. *Validated by PCR. **b**, Z-scores of normalized read depth of exome sequencing data around the *STRA8* locus, plotted against the *STRA8* gene model. NLS; nuclear localization; NES, nuclear export signal. **c**, Schematic representation of experimentally mapping the *STRA8* deletion in the carrier DNA. Three PCR reactions were used with primers anchoring in exons E3-E4 (PCR1, inside the predicted deletion region (pink box) spanning chr7:134,925,271-134,931,454, hg19) expected to be amplified in control (C) only and within exon E8 (PCR2, outside the deletion) amplified in both control and the patient. Third PCR reaction (PCR3) was designed to span the deletion region and enabled to determine the breakpoint-breakpoint junction of the deletion by Sanger sequencing. The deletion breakpoints were mapped at positions chr7:134925172 (BP1, 98 bp from E2) and chr7:134933449 (BP2, 2050 bp from E6). **d**, As expected, PCR1 did not yield a PCR product in the patient (P; the observed 800 bp band is a non-specific product), whereas PCR2 reaction was amplified in the patient. The PCR3 reaction spanning the deletion breakpoints resulted in a 5200 bp fragment in the patient when the expected size in control (C) was 13 kb. Neg, no DNA control; M, marker.

**Extended Data Fig. 6.**
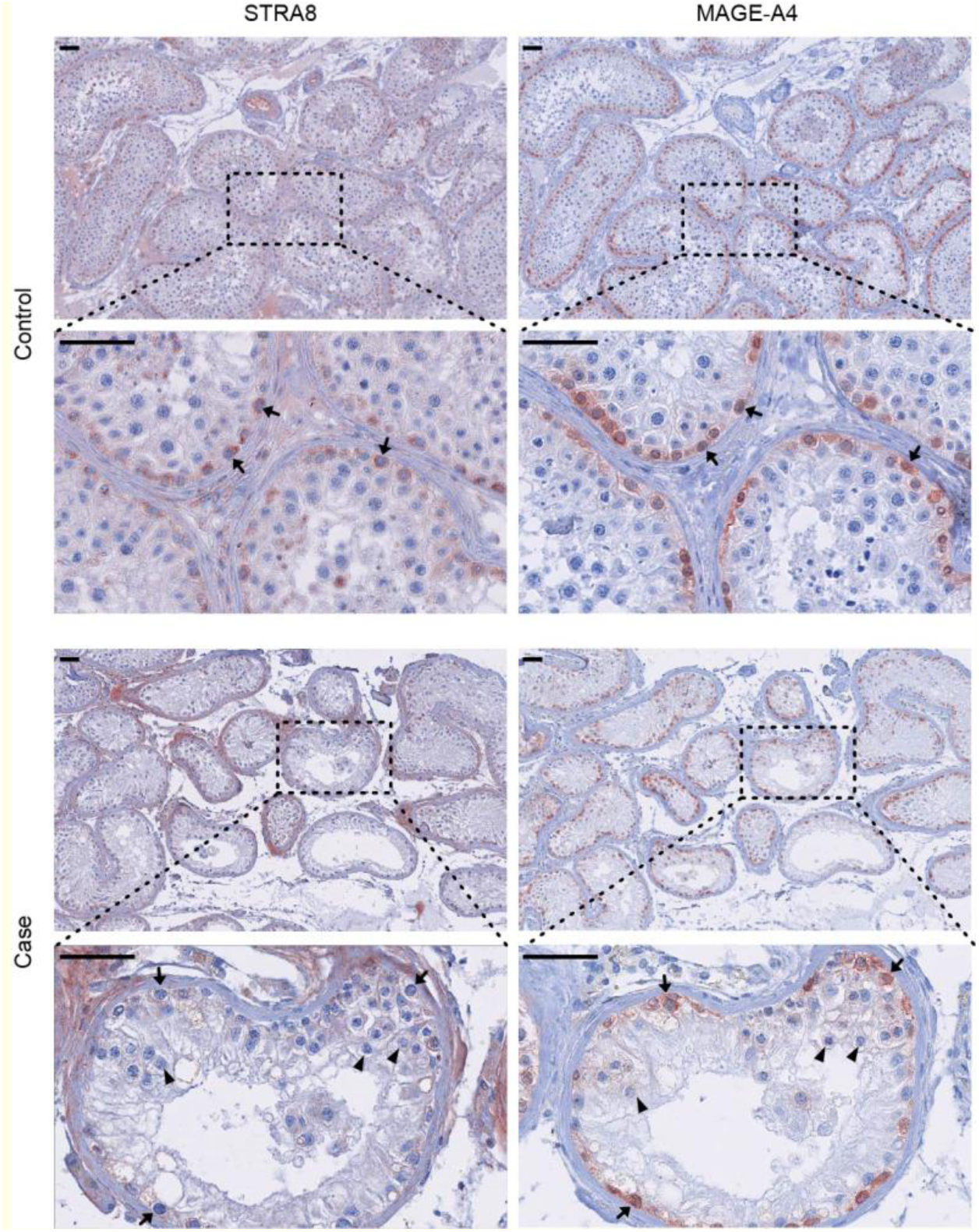
STRA8 and MAGE-A4 staining of testicular tissue. The carrier of *STRA8* deletion displayed significantly reduced staining of STRA8 in spermatogonia (arrows) compared to the control with normal spermatogenesis. The *STRA8* deletion caused a maturation arrest phenotype, whereby the tubules predominantly contained spermatogonia, stained with spermatogonia-specific marker MAGE-A4, and occasional pre-pachytene spermatocytes (arrowheads). Non-specific background staining is observed particularly in peritubular and Leydig cells in spite of extensive optimization of the only commercially available antibody.

**Extended Data Fig. 7.**
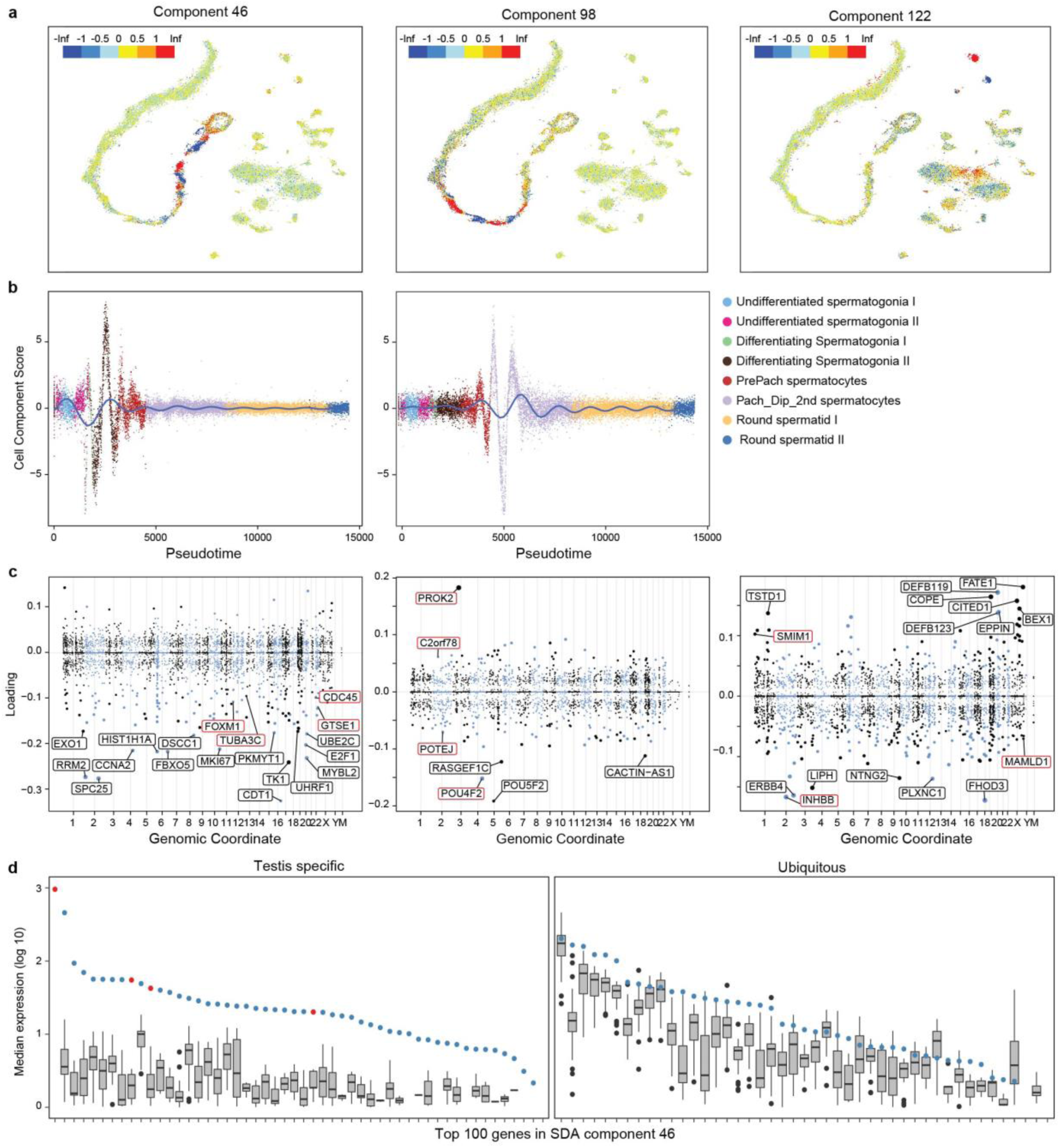
Examples of three potential subforms of azoospermia corresponding to defects in type B spermatogonia, spermatocytes, or Sertoli cells. **a**, Projection of cell loadings for the SDA components 46, 98 and 122 plotted in a t-SNE space. SDA component 46 appears to encode a set of genes specifically expressed in differentiating spermatogonia, whereas component 98 involves spermatocytes and component 122 involves Sertoli cells. tSNE plots use the same coordinates as in Figure 3 of the main text. **b**, Pseudotime (or germ cell development trajectory) plots for components 46 and 98. Note the primary cell type(s) expressing component 46 in a cycling manner, which probably correspond to the two mitotic divisions of Type B spermatogonia prior to meiosis. Like component 98 shows two cycles that may correspond to meiotic divisions. **c**, Manhattan plots showing the genes in the SDA components 46, 98 and 122, which can be used to infer the biological function(s) encoded by a component. Gene loadings corresponding to genes prioritized in the NOA cases are highlighted in red. **d**, Genes loading on component 46 can be divided into two groups; those with testis-enriched expression and those that are ubiquitous across the body. Each column corresponds to the expression pattern of a single component 46 gene: the box-and-whiskers plot summarizes gene expression across 52 tissues profiled by the GTEx project, while each point corresponds to the expression of that gene in testis alone. Red points correspond to genes identified as associated with male infertility in this study. Only expression levels above 1 transcripts per million (TPM) are visualized.

**Extended Data Fig. 8.**
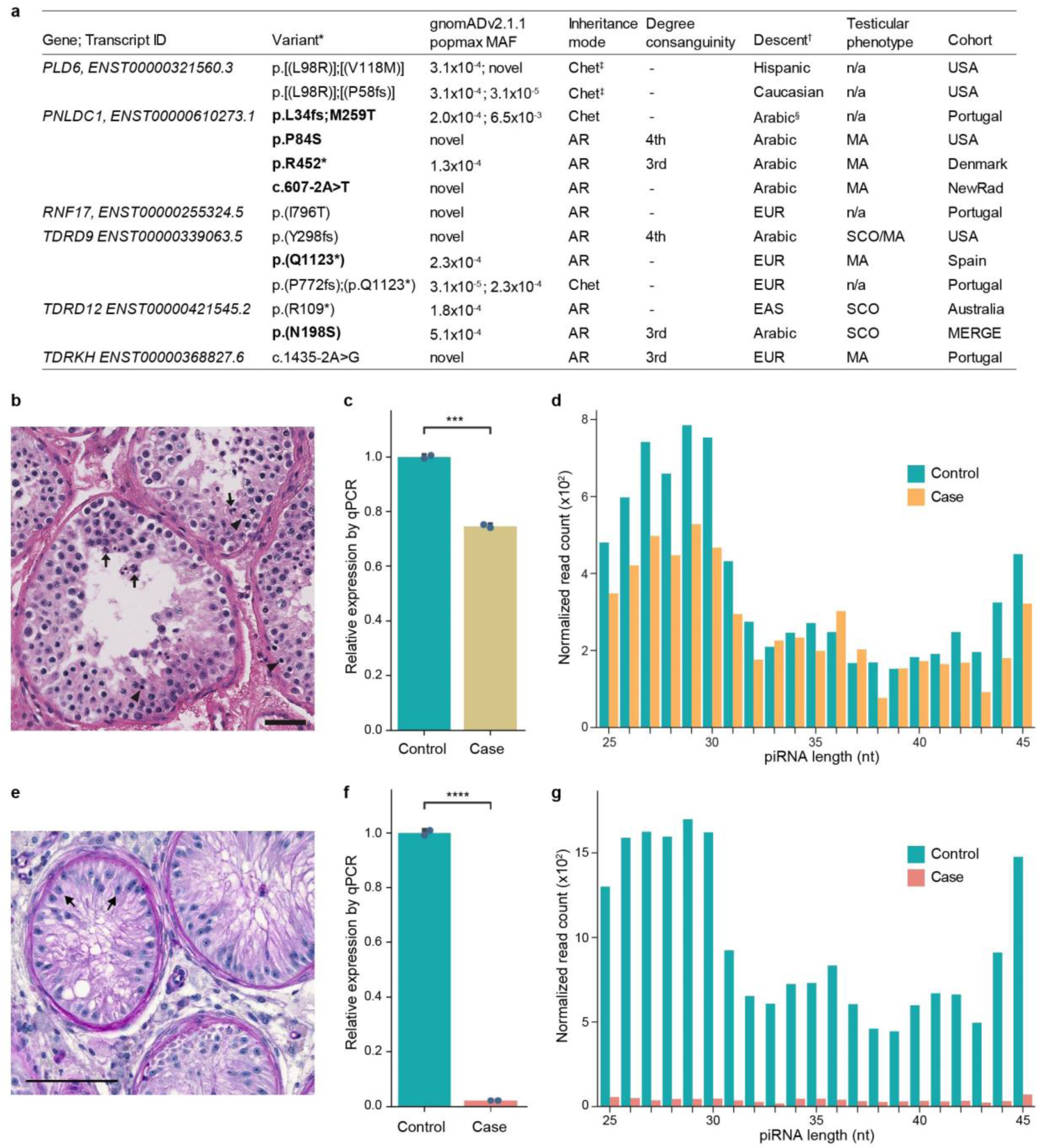
Characteristics and validation of prioritized variation detected in genes relevant for piRNA processing across male infertility cohorts. **a,** The characteristics of variation in piRNA processing genes. Carriers with testicular tissue available for experimental validation are indicated in bold. *Variant annotation based on ANNOVAR (Gencode v19), ^†^Self-reported descent or inferred ethnicity if no information available, ^‡^Compound heterozygous variation confirmed to be in trans configuration, ^§^Suggestive Arabic descent based on patient name. AR, autosomal recessive; Chet, compound heterozygous variation; NewRad, Newcastle/Radboud; SCO, Sertoli Cell Only; MA, maturation arrest; n/a, information unavailable. **b-d**, Validation of a rare stopgain variation p.(Q1123*) in the *TDRD9* gene. **b**, H&E stain of TDRD9 patient biopsy showing spermatogenic arrest. The most mature germ cell observed were early round spermatids, which often appeared multinucleated (arrows). In addition, many pyknotic cells were observed (arrowheads). Scale bar=50 *μ*m. **c**, The RT-qPCR analysis detected significantly reduced levels of the *TDRD9* transcript in the testicular tissue of the p.(Q1123*) carrier compared to the control with normal spermatogenesis (***P<0.001). **d**, Small RNA sequencing of the patient testicular tissue revealed reduced amounts of mature piRNA molecules (<32 nt) in the case compared to the control leading to a significant shift towards longer immature piRNAs (Fig. 5d). **e-g**, Analysis of the testicular tissue from the patient with the missense variation p.(N198S) in the *TDRD12* gene. **e**, The carrier of the p.(N198S) variation displayed Sertoli Cell Only syndrome in all seminiferous tubules, whereby only Sertoli cells are found (arrows) and no germ cells are observed. Testicular tissue was stained with Periodic Acid-Schiff (PAS) and the bar represents 100 µm. **f**, Relative expression of *TDRD12* demonstrated a nearly absent levels of the transcripts in the patient compared to the testicular tissue of a control with normal spermatogenesis (****P<0.0001). **g**, Size distribution of piRNAs detected in the NOA case with a biallelic missense variant in *TDRD12* and matching control, derived from small RNA-sequencing of testis tissue.

**Extended Data Table 1.** Fraction of individuals with prioritized rare SNVs and INDELs among NOA cases (n=924) and institutional controls (total, n=2265, n=851 men). Note the highest sensitivity of identifying a possible genetic cause of NOA in the autosomal homozygous inheritance mode, which was likely powered by the presence of consanguineous men in the NOA cohort. P-values below 0.05 were considered significant. *The sum of individuals across disease models is larger than “Total” as multiple candidate disease models can be observed per person.

EXTENDED DATA TABLES
| Disease model | NOA cases | Institutional controls |  |  |  |
| --- | --- | --- | --- | --- | --- |
|  | All (count) | All (count) | Fisher's exact P | Men (count) | Fisher's exact P |
| Single variant autosomal recessive | 11.0% (102) | 2.0% (46) | 7.0x10<sup>-25</sup> | 2.0% (17) | 2.4x10<sup>-15</sup> |
| Compound-heterozygote autosomal recessive | 7.3% (67) | 4.0% (91) | 2.1x10<sup>-4</sup> | 4.8% (41) | 3.7x10<sup>-2</sup> |
| X-linked hemizygote | 1.7% (16) | 0.2% (4) | 3.1x10<sup>-6</sup> | 0.5% (4) | 1.3x10<sup>-2</sup> |
| Y-linked hemizygote | 0.4% (4) | 0 | 7.0x10<sup>-3</sup> | 0 | 0.126 |
| Total* | 19.3% (178) | 6.0% (135) | 1.7x10<sup>-27</sup> | 6.9% (59) | 1.0x10<sup>-14</sup> |

**Extended Data Table 2.**
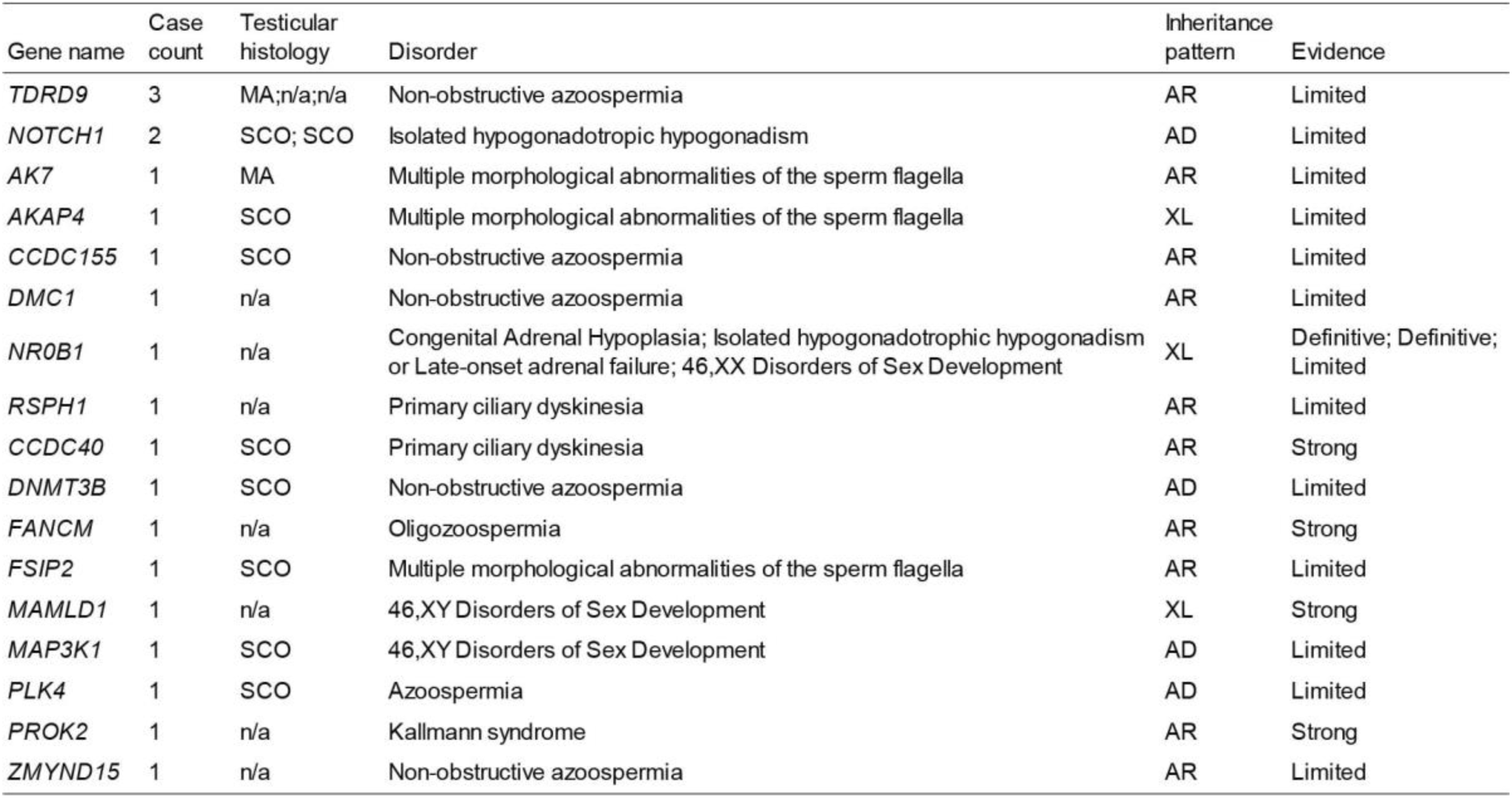
Known and suspected infertility genes (n=17) disrupted among NOA cases. List of genes linked to male infertility (n=164) was extracted from Oud et al. 2019^7^. Genes with ‘limited’ evidence should be considered as candidate genes that require additional proof of association. AD, Autosomal dominant, AR, Autosomal recessive, XL, X-linked, MA, Maturation arrest, SCO, Sertoli Cell Only, n/a, information unavailable.

| Gene name | Case count | Testicular histology | Disorder | Inheritance pattern | Evidence |
| --- | --- | --- | --- | --- | --- |
| *TDRD9* | 3 | MA; n/a; n/a | Non-obstructive azoospermia | AR | Limited |
| *NOTCH1* | 2 | SCO; SCO | Isolated hypogonadotropic hypogonadism | AD | Limited |
| *AK7* | 1 | MA | Multiple morphological abnormalities of the sperm flagella | AR | Limited |
| *AKAP4* | 1 | SCO | Multiple morphological abnormalities of the sperm flagella | XL | Limited |
| *CCDC155* | 1 | SCO | Non-obstructive azoospermia | AR | Limited |
| *DMC1* | 1 | n/a | Non-obstructive azoospermia | AR | Limited |
| *NR0B1* | 1 | n/a | Congenital Adrenal Hypoplasia; Isolated hypogonadotropic hypogonadism or Late-onset adrenal failure; 46,XX Disorders of Sex Development | XL | Definitive; Definitive; Limited |
| *RSPH1* | 1 | n/a | Primary ciliary dyskinesia | AR | Limited |
| *CCDC40* | 1 | SCO | Primary ciliary dyskinesia | AR | Strong |
| *DNMT3B* | 1 | SCO | Non-obstructive azoospermia | AD | Limited |
| *FANCM* | 1 | n/a | Oligozoospermia | AR | Strong |
| *FSIP2* | 1 | SCO | Multiple morphological abnormalities of the sperm flagella | AR | Limited |
| *MAMLD1* | 1 | n/a | 46,XY Disorders of Sex Development | XL | Strong |
| *MAP3K1* | 1 | SCO | 46,XY Disorders of Sex Development | AD | Limited |
| *PLK4* | 1 | SCO | Azoospermia | AD | Limited |
| *PROK2* | 1 | n/a | Kallmann syndrome | AR | Strong |
| *ZMYND15* | 1 | n/a | Non-obstructive azoospermia | AR | Limited |

## Notes

### Competing Interest Statement

The authors have declared no competing interest.

### Author Declarations

The study was approved by the Ethics Committee of all collaborative centers: protocols #201107177 and #201109261 approved by the institutional review board (IRB) of Washington University in St. Louis, USA; IRB_00063950 approved by IRB of University of Utah, USA; PTDC/SAU-GMG/101229/2008 approved by the Ethics Committee and Hospital Authority, University of Porto, Portugal; 16030459 and 0102004794 approved by the IRB of Weill Cornell Medical College, New York, USA; Ref. No.: 2014/04c approved by the IRB of Fundacio Puigvert, Barcelona, Spain; NL50495.091.14 version 4 approved by the Research Ethics Committee of the Radboud University, Nijmegen, The Netherlands; Ref: 18/NE/0089, Bursa: 05.01.2015/04 approved by the University Research Ethics Committee of University of Newcastle, UK; and ethical approvals from human ethics committees of Monash Surgical Day Hospital, Monash Medical Centre and Monash University, Australia; the ethics committee for the Capital Copenhagen Region (Ref. Nr. H-2-2014-103) and the Danish Personal Data Protection Agency (Datatilsynet 2012-58-0004, local Nr. 30-1482, I-Suite 03696) gave ethical approval for this work in accordance to the European Commission Directive for the transfer of personal data (MTA/I-4728.A1); the Ethics Committee of National Institute of Health Dr Ricardo Jorge, Lisboa, Portugal gave ethical approval for this work; and approval 74/54 (last amendment 288/M-13) released by Research Ethics Committee of the University of Tartu, Estonia. The MERGE study protocol was given ethical approval by the Ethics Committee of the Arztekammer Westfalen-Lippe and the University of Munster (Ref. No. 2010-578-f-S). Written informed consent was obtained from all men.

## REFERENCES

1. Jarow, J. P., Espeland, M. A. & Lipshultz, L. I. Evaluation of the Azoospermic Patient. Journal of Urology vol. 142 62–65 (1989).

2. Willott, G. M. Frequency of azoospermia. Forensic Sci. Int. 20, 9–10 (1982).

3. Itoh, N., Kayama, F., Tatsuki, T. J. & Tsukamoto, T. Have sperm counts deteriorated over the past 20 years in healthy, young Japanese men? Results from the Sapporo area. J. Androl. 22, 40–44 (2001).

4. Tüttelmann, F. et al. Clinical experience with azoospermia: aetiology and chances for spermatozoa detection upon biopsy. Int. J. Androl. 34, 291–298 (2011).

5. Giudice, F. D. et al. Clinical correlation among male infertility and overall male health: A systematic review of the literature. Investigative and Clinical Urology vol. 61 355 (2020).

6. Tiepolo, L. & Zuffardi, O. Localization of factors controlling spermatogenesis in the nonfluorescent portion of the human Y chromosome long arm. Hum. Genet. 34, 119–124 (1976).

7. Oud, M. S. et al. A systematic review and standardized clinical validity assessment of male infertility genes. Hum. Reprod. 34, 932–941 (2019).

8. Kasak, L. & Laan, M. Monogenic causes of non-obstructive azoospermia: challenges, established knowledge, limitations and perspectives. Hum. Genet. (2020) doi:10.1007/s00439-020-02112-y.

9. Nagirnaja, L., Aston, K. I. & Conrad, D. F. Genetic intersection of male infertility and cancer. Fertil. Steril. 109, 20–26 (2018).

10. Riera-Escamilla, A. et al. Sequencing of a ‘mouse azoospermia’ gene panel in azoospermic men: identification of RNF212 and STAG3 mutations as novel genetic causes of meiotic arrest. Human Reproduction vol. 34 978–988 (2019).

11. Fakhro, K. A. et al. Point-of-care whole-exome sequencing of idiopathic male infertility. Genet. Med. 20, 1365–1373 (2018).

12. Krausz, C. et al. Genetic dissection of spermatogenic arrest through exome analysis: clinical implications for the management of azoospermic men. Genet. Med. 22, 1956–1966 (2020).

13. Chen, S. et al. Whole-exome sequencing of a large Chinese azoospermia and severe oligospermia cohort identifies novel infertility causative variants and genes. Hum. Mol. Genet. 29, 2451–2459 (2020).

14. Zhang, H. et al. Whole exome sequencing identifies genes associated with non-obstructive azoospermia. doi:10.1101/2020.05.29.20116558.

15. Alhathal, N. et al. A genomics approach to male infertility. Genet. Med. 22, 1967–1975 (2020).

16. Wilfert, A. B. et al. Genome-wide significance testing of variation from single case exomes. Nat. Genet. 48, 1455–1461 (2016).

17. Tan, R. et al. An evaluation of copy number variation detection tools from whole-exome sequencing data. Hum. Mutat. 35, 899–907 (2014).

18. Lopes, A. M. et al. Human spermatogenic failure purges deleterious mutation load from the autosomes and both sex chromosomes, including the gene DMRT1. PLoS Genet. 9, e1003349 (2013).

19. Wyrwoll, M. J. et al. Bi-allelic Mutations in M1AP Are a Frequent Cause of Meiotic Arrest and Severely Impaired Spermatogenesis Leading to Male Infertility. Am. J. Hum. Genet. (2020) doi:10.1016/j.ajhg.2020.06.010.

20. Fang, K. et al. Prediction and Validation of Mouse Meiosis-Essential Genes Based on Spermatogenesis Proteome Dynamics. Mol. Cell. Proteomics 20, 100014 (2021).

21. Lim, E. T. et al. Rare Complete Knockouts in Humans: Population Distribution and Significant Role in Autism Spectrum Disorders. Neuron vol. 77 235–242 (2013).

22. Fon Tacer, K., et al. MAGE cancer-testis antigens protect the mammalian germline under environmental stress. Sci Adv 5, eaav4832 (2019).

23. Anderson, E. L. et al. Stra8 and its inducer, retinoic acid, regulate meiotic initiation in both spermatogenesis and oogenesis in mice. Proc. Natl. Acad. Sci. U. S. A. 105, 14976–14980 (2008).

24. Richards, S. et al. Standards and guidelines for the interpretation of sequence variants: a joint consensus recommendation of the American College of Medical Genetics and Genomics and the Association for Molecular Pathology. Genet. Med. 17, 405–424 (2015).

25. Djureinovic, D. et al. The human testis-specific proteome defined by transcriptomics and antibody-based profiling. Mol. Hum. Reprod. 20, 476–488 (2014).

26. Consortium, T. G. & The GTEx Consortium. The GTEx Consortium atlas of genetic regulatory effects across human tissues. Science vol. 369 1318–1330 (2020).

27. Mahyari, E. et al. Comparative single-cell analysis of biopsies clarifies pathogenic mechanisms in Klinefelter syndrome. Am. J. Hum. Genet. 108, 1924–1945 (2021).

28. Jung, M. et al. Unified single-cell analysis of testis gene regulation and pathology in five mouse strains. Elife 8, (2019).

29. Ehmcke, J. & Schlatt, S. A revised model for spermatogonial expansion in man: lessons from non-human primates. Reproduction 132, 673–680 (2006).

30. Hess, R. A. & Renato de Franca, L. Spermatogenesis and cycle of the seminiferous epithelium. Adv. Exp. Med. Biol. 636, 1–15 (2008).

31. Anderson, R. A. & Sharpe, R. M. Regulation of inhibin production in the human male and its clinical applications. Int. J. Androl. 23, 136–144 (2000).

32. Ogata, T., Sano, S., Nagata, E., Kato, F. & Fukami, M. MAMLD1 and 46,XY disorders of sex development. Semin. Reprod. Med. 30, 410–416 (2012).

33. Ernst, C., Odom, D. T. & Kutter, C. The emergence of piRNAs against transposon invasion to preserve mammalian genome integrity. Nat. Commun. 8, 1411 (2017).

34. Fu, Q. & Wang, P. J. Mammalian piRNAs: Biogenesis, function, and mysteries. Spermatogenesis 4, e27889 (2014).

35. Ding, D. et al. PNLDC1 is essential for piRNA 3’ end trimming and transposon silencing during spermatogenesis in mice. Nat. Commun. 8, 819 (2017).

36. Huang, H. et al. piRNA-associated germline nuage formation and spermatogenesis require MitoPLD profusogenic mitochondrial-surface lipid signaling. Dev. Cell 20, 376–387 (2011).

37. Watanabe, T. et al. MITOPLD is a mitochondrial protein essential for nuage formation and piRNA biogenesis in the mouse germline. Dev. Cell 20, 364–375 (2011).

38. Wasik, K. A. et al. RNF17 blocks promiscuous activity of PIWI proteins in mouse testes. Genes Dev. 29, 1403–1415 (2015).

39. Saxe, J. P., Chen, M., Zhao, H. & Lin, H. Tdrkh is essential for spermatogenesis and participates in primary piRNA biogenesis in the germline. EMBO J. 32, 1869–1885 (2013).

40. Shoji, M. et al. The TDRD9-MIWI2 complex is essential for piRNA-mediated retrotransposon silencing in the mouse male germline. Dev. Cell 17, 775–787 (2009).

41. Pandey, R. R. et al. Tudor domain containing 12 (TDRD12) is essential for secondary PIWI interacting RNA biogenesis in mice. Proc. Natl. Acad. Sci. U. S. A. 110, 16492–16497 (2013).

42. Arafat, M. et al. Mutation in TDRD9 causes non-obstructive azoospermia in infertile men. J. Med. Genet. 54, 633–639 (2017).

43. Li, Z. et al. Excess of rare variants in genes that are key epigenetic regulators of spermatogenesis in the patients with non-obstructive azoospermia. Sci. Rep. 5, 8785 (2015).

44. Girard, A., Sachidanandam, R., Hannon, G. J. & Carmell, M. A. A germline-specific class of small RNAs binds mammalian Piwi proteins. Nature 442, 199–202 (2006).

45. Nagirnaja, L. et al. Variant, Defective piRNA Processing, and Azoospermia. N. Engl. J. Med. 385, 707–719 (2021).

46. Gunes, S., Arslan, M. A., Hekim, G. N. T. & Asci, R. The role of epigenetics in idiopathic male infertility. J. Assist. Reprod. Genet. 33, 553–569 (2016).

47. Oud, M. S. et al. A de novo paradigm for male infertility. Nat. Commun. 13, 154 (2022).

48. DiStefano, M. T. et al. ClinGen expert clinical validity curation of 164 hearing loss gene-disease pairs. Genet. Med. 21, 2239–2247 (2019).

49. Duncan, J. L. et al. Inherited Retinal Degenerations: Current Landscape and Knowledge Gaps. Transl. Vis. Sci. Technol. 7, 6 (2018).

50. Wheway, G., Mitchison, H. M. & Genomics England Research Consortium. Opportunities and Challenges for Molecular Understanding of Ciliopathies–The 100,000 Genomes Project. Frontiers in Genetics vol. 10 (2019).

51. Thormann, A. et al. Flexible and scalable diagnostic filtering of genomic variants using G2P with Ensembl VEP. Nat. Commun. 10, 2373 (2019).

52. Kousi, M. & Katsanis, N. Genetic modifiers and oligogenic inheritance. Cold Spring Harb. Perspect. Med. 5, (2015).

53. Chivukula, R. R. et al. A human ciliopathy reveals essential functions for NEK10 in airway mucociliary clearance. Nat. Med. 26, 244–251 (2020).

54. Dong, F. N. et al. Absence of CFAP69 Causes Male Infertility due to Multiple Morphological Abnormalities of the Flagella in Human and Mouse. Am. J. Hum. Genet. 102, 636–648 (2018).

55. Tang, S. et al. Biallelic Mutations in CFAP43 and CFAP44 Cause Male Infertility with Multiple Morphological Abnormalities of the Sperm Flagella. Am. J. Hum. Genet. 100, 854–864 (2017).

56. Martinez, G. et al. Whole-exome sequencing identifies mutations in FSIP2 as a recurrent cause of multiple morphological abnormalities of the sperm flagella. Hum. Reprod. 33, 1973–1984 (2018).

57. Rozen, S. G. et al. AZFc deletions and spermatogenic failure: a population-based survey of 20,000 Y chromosomes. Am. J. Hum. Genet. 91, 890–896 (2012).

58. Zoch, A. et al. SPOCD1 is an essential executor of piRNA-directed de novo DNA methylation. Nature 584, 635–639 (2020).

59. Huang, L., Ma, F., Chapman, A., Lu, S. & Xie, X. S. Single-Cell Whole-Genome Amplification and Sequencing: Methodology and Applications. Annu. Rev. Genomics Hum. Genet. 16, 79–102 (2015).

60. Kang, E. et al. Mitochondrial replacement in human oocytes carrying pathogenic mitochondrial DNA mutations. Nature 540, 270–275 (2016).

